# Epidemiology of scrub typhus and other rickettsial infections (2018-22) in the hyper-endemic setting of Mizoram, North-East India

**DOI:** 10.1101/2023.04.21.23288926

**Authors:** Vanramliana, Lalfakzuala Pautu, Pachuau Lalmalsawma, Gabriel Rosangkima, Devojit Kumar Sarma, Hunropuia, Yogesh Malvi, Naveen Kumar Kodali, Christiana Amarthaluri, K Balasubramani, Praveen Balabaskaran Nina

## Abstract

In the last decade, there has been an emergence of scrub typhus in many parts of India. In Mizoram, North-East India, there has been a steep increase in scrub typhus and other rickettsial infections in the last 5 years. As part of the public health response, the Mizoram Government has integrated screening (by rapid immunochromatographic test and/or Weil-Felix test) and line listing of scrub typhus and other rickettsial infections across all its health settings, a first in India. From 2018-22 (study period), 22914 cases were reported; of these, 19651 were positive for scrub typhus. Aizawl district is the worst affected, with 10580 cases (46.17%). The average incidence rate of rickettsial infections is 3.54 cases per 1000 persons-year, and the case fatality rate is 0.35. Patients with eschar (aOR=2.5, p<0.05), construction workers (aOR=17.9, p<0.05), and children aged 10 and below (aOR=5.4, p<0.05) have higher odds of death due to rickettsial infections.

---

Scrub typhus is a re-emerging infectious zoonotic disease caused by *Orientia tsutsugamushi*, an obligatory, gram-negative, intracellular bacterium; the bite of an infected larval chigger containing *O. tsutsugamushi* leads to human infection [1]. Due to its high morbidity and mortality, scrub typhus infection is considered to be serious and life-threatening [2]; the case fatality rate (CFR) can reach as high as 70% if untreated [3]. Scrub typhus mortality is influenced by the virulence of the strains, patient characteristics, delay in diagnosis, and drug resistance [3, 4].

Eschar, the first pathognomonic lesion and a key diagnostic marker [5] of scrub typhus, is often seen at the site of the chigger bite [6]. Eschar often goes unnoticed, as the vector bite is painless, despite being pruritic on occasion [7]. However, the presence or absence of eschar does not alter the severity of the disease outcome [3]. From the eschar stage, the infection advances to an acute febrile illness [8] with varied clinical presentations such as fever, myalgia, nausea, rash, lymphadenopathy, vomiting, diarrhoea, cough, and breathlessness [9]. Mostly in untreated cases, the disease progresses to different organs and organ systems [10, 11], and in some cases, is clinically manifested as multi-organ dysfunction syndrome [6].

In addition to scrub typhus, serosurveys carried out in scrub typhus endemic regions have also revealed the endemicity of other rickettsial infections caused by the spotted fever group (SFG)—(Epidemic louse borne typhus and Endemic typhus), and typhus group (TG)—(Rocky Mountain spotted fever, Rickettsialpox, and Indian tick typhus) rickettsiae [12-14]; these infections share almost similar signs and symptoms with scrub typhus [14]. Just like scrub typhus, other rickettsial infections are associated with significant mortality unless diagnosed and treated early [15].

In India, after a gap of 65 years, scrub typhus is re-emerging in different regions [16]. Mizoram, the landlocked North-East Indian state bordering Myanmar and Bangladesh to its east and west, respectively is the worst affected [17, 18]. In the last five years (2018-2022), Mizoram has seen a substantial increase in scrub typhus incidence; Mizoram’s 22914 cases in the last five years are higher than rest of India’s decadal (2010-2020) cumulative count (18781) [19]. With a forest cover of 88.48%, Mizoram is a biodiversity hotspot, and 60% of its population is engaged in agricultural practices [20]. A recent seroprevalence survey reports 46% of the rodents in Mizoram to be positive for scrub typhus antibodies, and 12 scrub typhus outbreaks across Mizoram from 2015-19 [21].

Given the rapid spread of scrub typhus and other rickettsial infections in the state, the Mizoram Government integrated screening and line listing of scrub typhus and other rickettsial infections from 2018 onwards across all its health centers. Here, we report the first comprehensive epidemiological study of scrub typhus and other rickettsial infections from 2018-2022, systematically recorded across a state in India.

## Methods

### Data source

Five years line listing data of scrub typhus and other rickettsial infections from 2018-2022 were obtained from Integrated Disease Surveillance Program (IDSP) under Health & Family Welfare Department, Government of Mizoram (Supplementary File 1). The line listing format includes information on name, age, sex, address, occupation, general symptoms, presence/absence of eschar, date of diagnosis, diagnostic kit utilized, hospitalized or not and outcome (recovered/expired). The line listing data were collected from all Government primary health centers (PHCs), community health centers (CHCs), district hospitals (DHs) and private hospitals. Until 2018, screening was mostly done using the scrub typhus specific Immunochromatographic test (ICT) - InBios rapid test (SD Bioline Tsutsugamushi test, SD Diagnostics, Hagal-dong, Kyonggi-do, Korea) [22]. However, as there were widespread reports of scrub typhus-like illness which were testing negative with the ICT across Mizoram, Government of Mizoram introduced and distributed the Weil-Felix (WF) test kits to all its testing units from 2019 onwards; reaction with OXK antigen is suggestive of scrub typhus, whereas reaction with OX19 and OX2 antigens suggests infection by TG and SFG, respectively [23, 24]. Based on WF positivity, rickettsial infections were classified into scrub typhus (OXK), other rickettsial infections (OX2, OX19, OX2 and OX19), mixed infections of scrub typhus with other rickettsiae (OXK and OX2, OXK, and OX19, and OXK, OX2, and OX19).

### Statistics

Based on the line listing data of rickettsial infections from 2018 to 2022, a preliminary analysis, including frequency tables of the recorded socio-demographic and clinical variables and their association with rickettsial infections (chi-square test) was carried-out using the IBM statistical software, SPSS (version 16.0). The chi-square analysis was also carried-out to determine the association between eschar and scrub typhus outcome.

A multivariate logistic regression analysis was run to estimate the association of demographic (age, sex, place of residence, occupation) and clinical (eschar and hospitalization) variables with the disease outcome (death or recovery). Age was grouped into 5 categories (< 11 years, 11-30 years, 31-50 years, 51-70 years and 71 and more). Districts were categorized into West (Mamit, Lunglei, Lawngtlai, Kolasib), Central (Aizawl, Serchhip) and East (Champhai, Khawzawl, Saitual, Hnahthial, Siaha). There was no issue of multicollinearity between independent variables. Adjusted odds ratios (aOR) and unadjusted odds ratios (uOR) with a confidence interval (CI) of 95% were calculated; p-value < 0.05 is considered significant. Incidence rates for rickettsial infections were calculated per 1000 persons [25] and were computed by retrieving dataset of the projected population estimates, 2020 [26].

### Spatial mapping and GIS analysis

To understand the spatial distribution of rickettsial infections, the patients’ addresses in the line listing data were converted into a spatial database. After the data cleaning process, the addresses of scrub typhus cases were searched in the Google map database for location information. A total of 836 case locations were identified and their corresponding latitudes and longitudes were extracted. The extracted locations were assigned a unique spatial ID and joined with the case details to prepare a spatial database using the ArcGIS 10.4 software (https://desktop.arcgis.com). The total cases recorded in each case location from 2018-2022 were used to identify hotspots of scrub typhus in Mizoram using the Optimized Hotspot Analysis tool in the ArcGIS Pro 3.0 software, as detailed [27, 28]. The size of the dots is proportionate to the total cumulative cases in each location. The hotspots were prepared as dot distribution map to represent the clusters of rickettsial infections with multiple significant (90%, 95%, and 99% confidence intervals) and non-significant levels.

## Results

### Incidence and distribution of rickettsial infections across the different districts of Mizoram

In Mizoram, from 2018-2022, 22914 cases of rickettsial infections, reactive for either the Tsutsugamushi test or the Weil-Felix test were reported from the eleven districts in the state. Among the 22914 cases, 19651 were considered to be scrub typhus based on Tsutsugamushi 1or Weil-Felix OXK positivity. The maximum number of scrub typhus cases were reported from Aizawl district (9863 cases, 50.19%), and was followed by Serchhip (2975 cases, 15.14%), while Champhai (2.07%) and Khawzawl (2.31%) reported the lowest (Table 1, Figure 1). The cases increased from 2018 to 2019 and were followed by a decline in 2020 and 2021. However, in 2022, there was an upsurge in scrub typhus (6542 cases), the highest in the past five years (Supplementary Table 1, Figure 1). Supplementary Table 2 details the district-wise distribution of different rickettsial infections (scrub typhus (OXK), other rickettsial infections (OX2, OX19, OX2 and OX19), mixed infections of scrub typhus with other rickettsiae (OXK and OX2, OXK, and OX19, and OXK, OX2, and OX19) diagnosed by the WF test, from 2019 to 2022. Over the study period, the scrub typhus caseload has increased substantially; from 297 cases in 2019, it has increased more than 13 times to 3932 cases in 2022 (Supplementary Table 2). A fourfold increase in mixed infections is also seen from 2019 (141 cases) to 2022 (562); by 2022, all the districts have reported mixed infections (Supplementary Table 2). The caseload (472) of other rickettsial infections (OX2, OX19, and OX2 and OX19 reactive), though higher than scrub typhus in 2019 (297), was outnumbered by scrub typhus cases in the following years (2020-22) (Supplementary Table 2). During the study period, the trend of other rickettsial infections was inconsistent; these are higher in the districts of Champhai, Hnahthial, Khawzawl, Kolasib, Mamit and Lunglei (Supplementary Table 2).

**Table 1.** Socio-demographic and clinical profile of rickettsial infections in Mizoram (2018-2022)

|  | Rickettsial infections |  |  | Total | p-value |
| --- | --- | --- | --- | --- | --- |
|  | Scrub typhus* | Other rickettsial infections<sup>†</sup> | Mixed (scrub typhus and other rickettsial) infections <sup>‡</sup> |  |  |
|  | 19651 (85.76%) | 1846 (8.06%) | 1417 (6.18%) | 22914 (100%) |  |
| Age group (in Years) |  |  |  |  |  |
| Less than 11 | 2374 (88.52) | 168 (6.26) | 140 (5.22) | 2682 (11.70) | 0.000 |
| 11 - 30 | 4830 (86.62) | 422 (7.57) | 324 (5.81) | 5576 (24.33) |  |
| 31 - 50 | 6731 (85.33) | 673 (8.53) | 484 (6.14) | 7888 (34.42) |  |
| 51 - 70 | 4421 (85.02) | 418 (8.04) | 361 (6.94) | 5200 (22.69) |  |
| 71 and above | 1295 (82.59) | 165 (10.52) | 108 (6.89) | 1568 (6.84) |  |
| Occupation |  |  |  |  |  |
| Farmer | 9805 (84.35) | 1048 (9.02) | 771 (6.63) | 11624 (50.73) | 0.000 |
| Business | 1955 (89.35) | 109 (4.98) | 124 (5.67) | 2188 (9.55) |  |
| Construction workers | 1104 (85.91) | 111 (8.64) | 70 (5.45) | 1285 (5.61) |  |
| Government services | 1344 (86.71) | 115 (7.42) | 91 (5.87) | 1550 (6.76) |  |
| Pre school | 968 (89.71) | 47 (4.36) | 64 (5.93) | 1079 (4.71 ) |  |
| Student | 4301 (86.33) | 402 (8.07) | 279 (5.60) | 4982 (21.74) |  |
| Others (aged 71 and above) | 174 (84.47) | 14 (6.80) | 18 (8.74) | 206 (0.90) |  |
| Sex |  |  |  |  |  |
| Male | 10278 (86.45) | 890 (7.49) | 721 (6.06) | 11889 (51.89) | 0.003 |
| Female | 9373 (85.02) | 956 (8.67) | 696 (6.31) | 11025 (48.11) |  |
| Districts |  |  |  |  |  |
| Aizawl | 9863 (93.22) | 149 (1.41) | 568 (5.37) | 10580 (46.17) | 0.000 |
| Champhai | 407 (67.72) | 164 (27.29) | 30 (4.99) | 601 (2.62) |  |
| Hnahthial | 531 (53.69) | 350 (35.39) | 108 (10.92) | 989 (4.32) |  |
| Khawzawl | 453 (46.18) | 452 (46.08) | 76 (7.75) | 981 (4.28) |  |
| Kolasib | 459 (61.45) | 191 (25.57) | 97 (12.99) | 747 (3.26) |  |
| Lawngtlai | 1357 (86.82) | 10 (0.64) | 196 (12.54) | 1563 (6.82) |  |
| Lunglei | 1125 (87.41) | 143 (11.11) | 19 (1.48) | 1287 (5.62) |  |
| Mamit | 1082 (72.42) | 236 (15.80) | 176 (11.78) | 1494 (6.52) |  |
| Saitual | 894 (85.71) | 83 (7.96) | 66 (6.33) | 1043 (4.55) |  |
| Serchhip | 2975 (95.60) | 57 (1.83) | 80 (2.57) | 3112 (13.58) |  |
| Siaha | 505 (97.68) | 11 (2.13) | 1 (0.19) | 517 (2.26) |  |
| Hospitalization |  |  |  |  |  |
| Yes | 3832 (91.63) | 148 (3.54) | 202 (4.83) | 4182 (18.25) | 0.000 |
| No | 15819 (84.45) | 1698 (9.06) | 1215 (6.49) | 18732 (81.75) |  |
| Presence of eschar |  |  |  |  |  |
| Yes | 443 (76.25) | 92 (15.83) | 46 (7.92) | 581 (2.54) | 0.000 |
| No | 19208 (86.01) | 1754 (7.85) | 1371 (6.14) | 22333 (97.46) |  |

p<0.05 is significant \*Scrub typhus: Rapid tsutsugamushi and/or WF OXK positive <sup>†</sup>Other rickettsial infections: WF OX2 and/or OX19 positive <sup>‡</sup>Mixed infections: WF OXK with OX2 and/or OX19 positive

**Figure 1.**
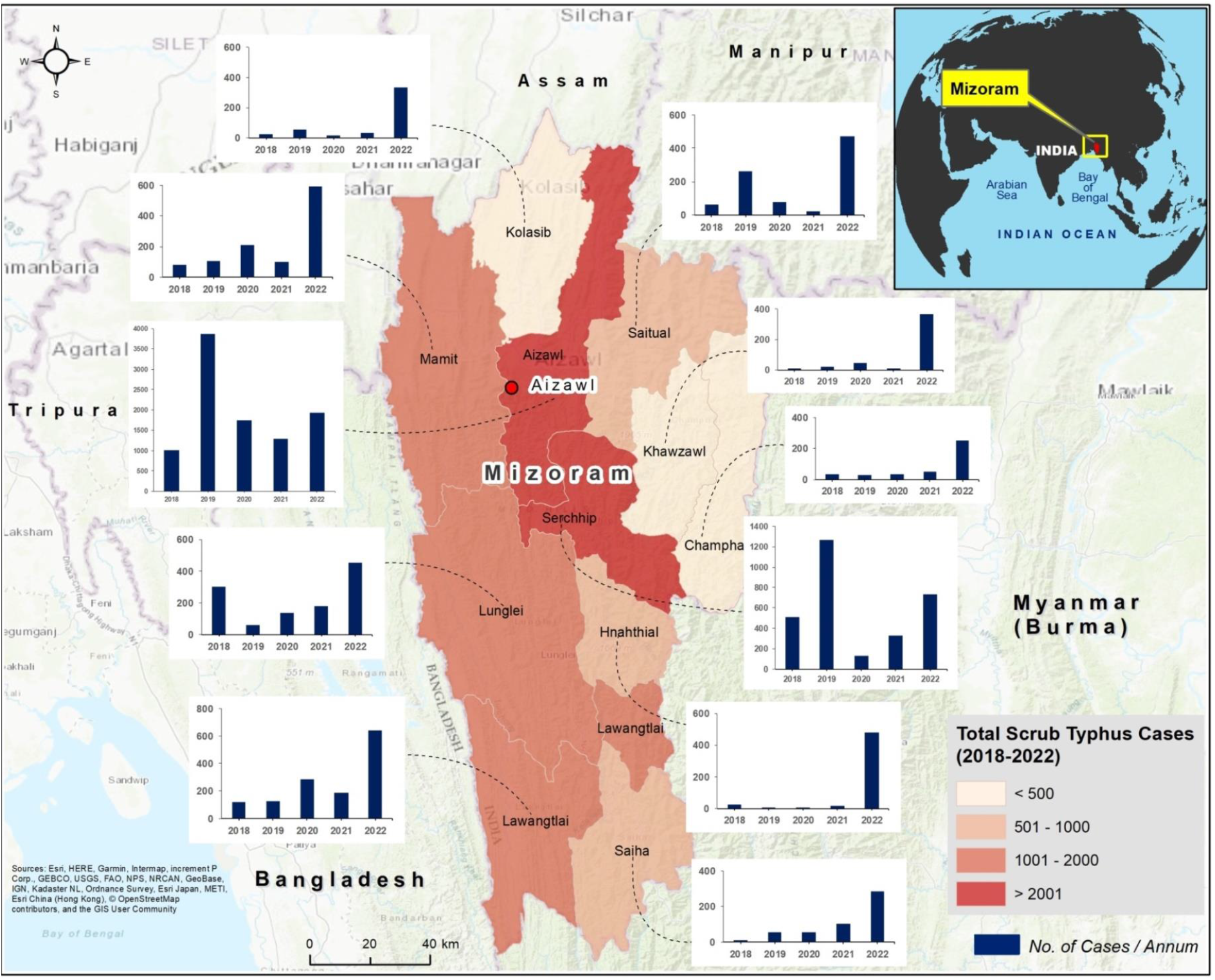
Distribution of scrub typhus cases across the districts of Mizoram (2018-2022). The trends in case distribution are represented as located bar chart. The y-axis of the graph shows the total cases in each year from 2018-2022. The length of the y-axis varies according to the caseloads across the districts of Mizoram. The background base map represents topography. For more information about the base map services, visit http://goto.arcgisonline.com/maps/World_Topo_Map

The incidence rates of rickettsial infections (scrub typhus, other rickettsial infections, and mixed infections) are shown in Table 2 and Supplementary Tables 3, 4; as scrub typhus constitutes the majority of infections, the incidence rate for scrub typhus and rickettsial infections are similar. The average incidence rate of scrub typhus cases in Mizoram from 2018-2022 is 3.04 cases per 1000. Serchhip’s average incidence rate (9.34) is much higher than the state’s average (3.02) and is almost double that of Aizawl (4.32) (Supplementary Table 3). Besides Serchhip and Aizawl, Saitual reported a slightly higher incidence rate (3.12) than the state’s average. Among all the districts, Champhai reported the least incidence of scrub typhus (0.79 per 1000) (Supplementary Table 3). The districts of Hnahthial and Khawzawl, have reported higher incidences of other rickettsial infections, with average incidence rates of 2.45 and 3.03 per 1000 persons/year, respectively (Supplementary Table 3). The incidence of mixed infections (scrub typhus and other rickettsial infections) was relatively less during 2019-2021, however, in 2022, a notable rise in the incidence rates was observed across Hnahthial (1.93), Khawzawl (1.45), Serchhip (1.19), and Mamit (1.06) (Supplementary Table 3). The incidence rate of OX2 was high in Hnahthial (2.17), while for OX19, it was Khawzawl (1.83) (Supplementary Table 4).

**Table 2.** Incidence rates of rickettsial infections across the districts of Mizoram (2018-2022)

| Cases per<br>1000<br>persons-year | Aizawl | Champhai | Hnahthial | Khawzawl | Kolasib | Lawngtlai | Lunglei | Mamit | Saitual | Serchhip | Siaha | Mizoram |
| --- | --- | --- | --- | --- | --- | --- | --- | --- | --- | --- | --- | --- |
| Rickettsial<br>infections* |  |  |  |  |  |  |  |  |  |  |  |  |
| 2018 | 2.21 | 0.33 | 0.76 | 0.24 | 0.21 | 0.80 | 2.17 | 0.80 | 1.08 | 8.05 | 0.17 | 1.69 |
| 2019 | 8.69 | 0.96 | 3.55 | 7.38 | 0.89 | 0.95 | 0.43 | 1.20 | 4.53 | 19.88 | 1.23 | 5.00 |
| 2020 | 4.30 | 0.80 | 4.36 | 5.45 | 1.05 | 2.26 | 1.50 | 3.90 | 1.38 | 2.10 | 1.27 | 2.87 |
| 2021 | 3.39 | 0.99 | 2.32 | 0.70 | 1.19 | 1.93 | 1.70 | 1.66 | 1.61 | 5.51 | 2.20 | 2.41 |
| 2022 | 4.56 | 2.72 | 16.67 | 12.56 | 3.54 | 4.62 | 3.51 | 7.69 | 9.63 | 13.31 | 6.08 | 5.73 |
| Avg.<br>incidence<br>rate | 4.63 | 1.16 | 5.53 | 5.27 | 1.38 | 2.11 | 1.86 | 3.05 | 3.65 | 9.77 | 2.19 | 3.54 |

\*Positive by rapid tsutsugamushi and/or WF (OXK/OX2/OX19) test

The spatial distribution shows the scrub typhus cases are predominately distributed in the populated (North-Central) districts, especially along the arterial route between Aizawl and Serchhip (Figure 2a). Even though urban centers have higher case locations, loci with >100 cumulative cases are generally distributed in the urban outskirts (Figure 2a). Scrub typhus hotspots in Mizoram are shown in Figure 2b. Most of the settlements in the Saitual district are primary hotspots of scrub typhus (99% significant). The primary hotspots also extended into the Aizawl district, especially along the borders of Saitual and Aizawl districts. The capital city of Mizoram, Aizawl and its neighbouring major town Serchhip are secondary hotspots for rickettsial infections. Siaha, Lawangtlai and Lunglei districts are coldspots of scrub typhus; these locations have less than 25 total cases during 2018-2022.

**Figure 2.**
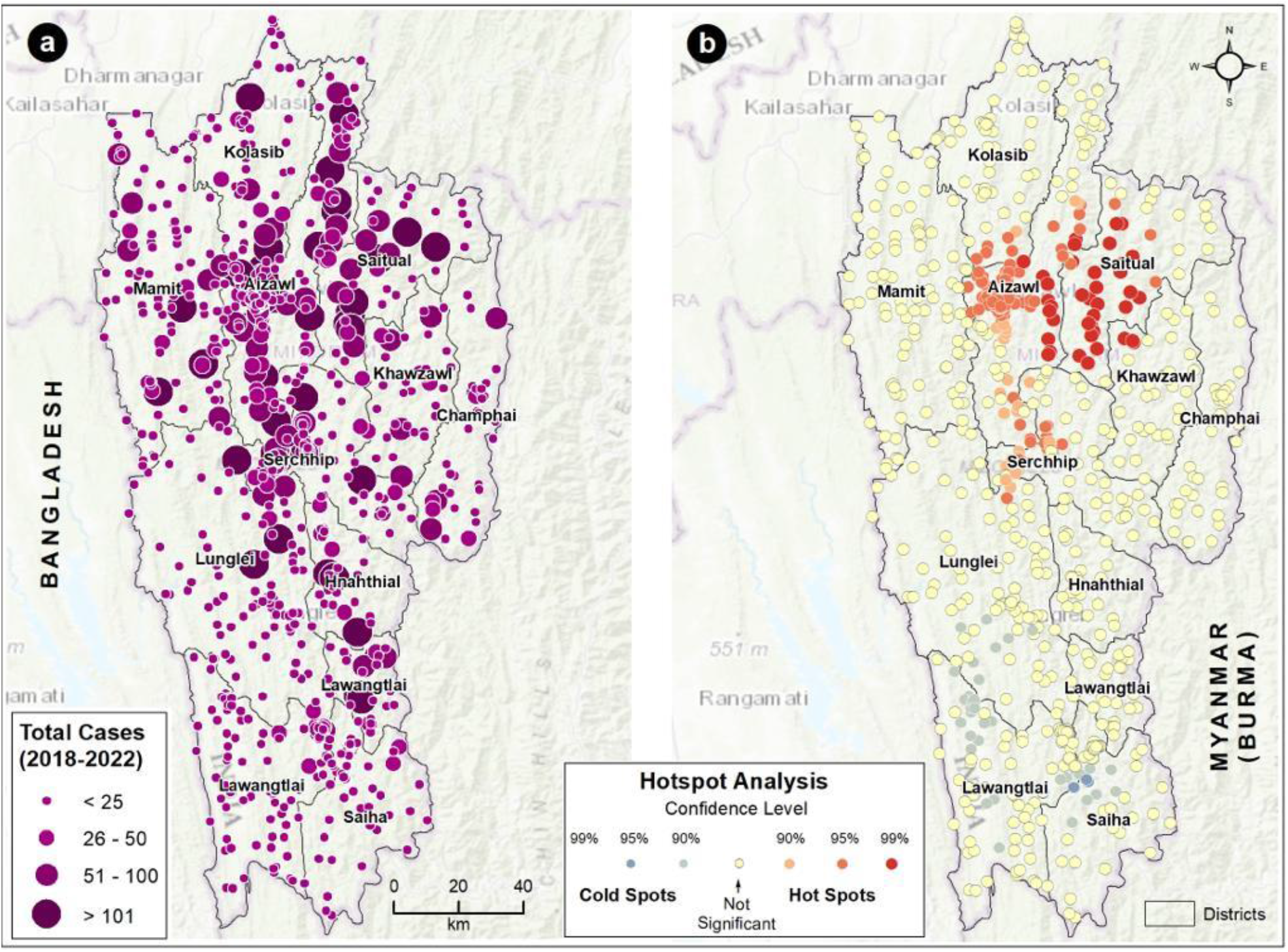
Distribution of (a) scrub typhus cases (2018-2022) and (b) hotspots in Mizoram. The large dot with dark purple color (a) indicates higher caseloads (>101). The color (purple) intensity and size of the dot gradually decrease with a reduction in the caseload. The red and blue shaded dots represent (b) the hot and cold spots, respectively, of scrub typhus cases in Mizoram with different statistical significance. The yellow-colored dots are not statistically significant for forming hot or cold spots. The background base map represents topography. For more information about the base map services, visit http://goto.arcgisonline.com/maps/World_Topo_Map

### Seasonal distribution of scrub typhus cases in Mizoram

A spike in scrub typhus cases was observed during the monsoon and post-monsoon seasons (June to November) (Figure 3). Together, monsoon and post-monsoon months account for more than 60% of the cases. July accounted for the maximum number of cases (2641, 13.44%), followed by August (2494, 12.7%). Aside from monsoon and post-monsoon months, scrub typhus cases were reported more in January (1794, 9.13%). Cases were also reported during the summer (March to May), accounting for 17.12% of the total cases (Figure 3, Supplementary Table 1).

**Figure 3.**
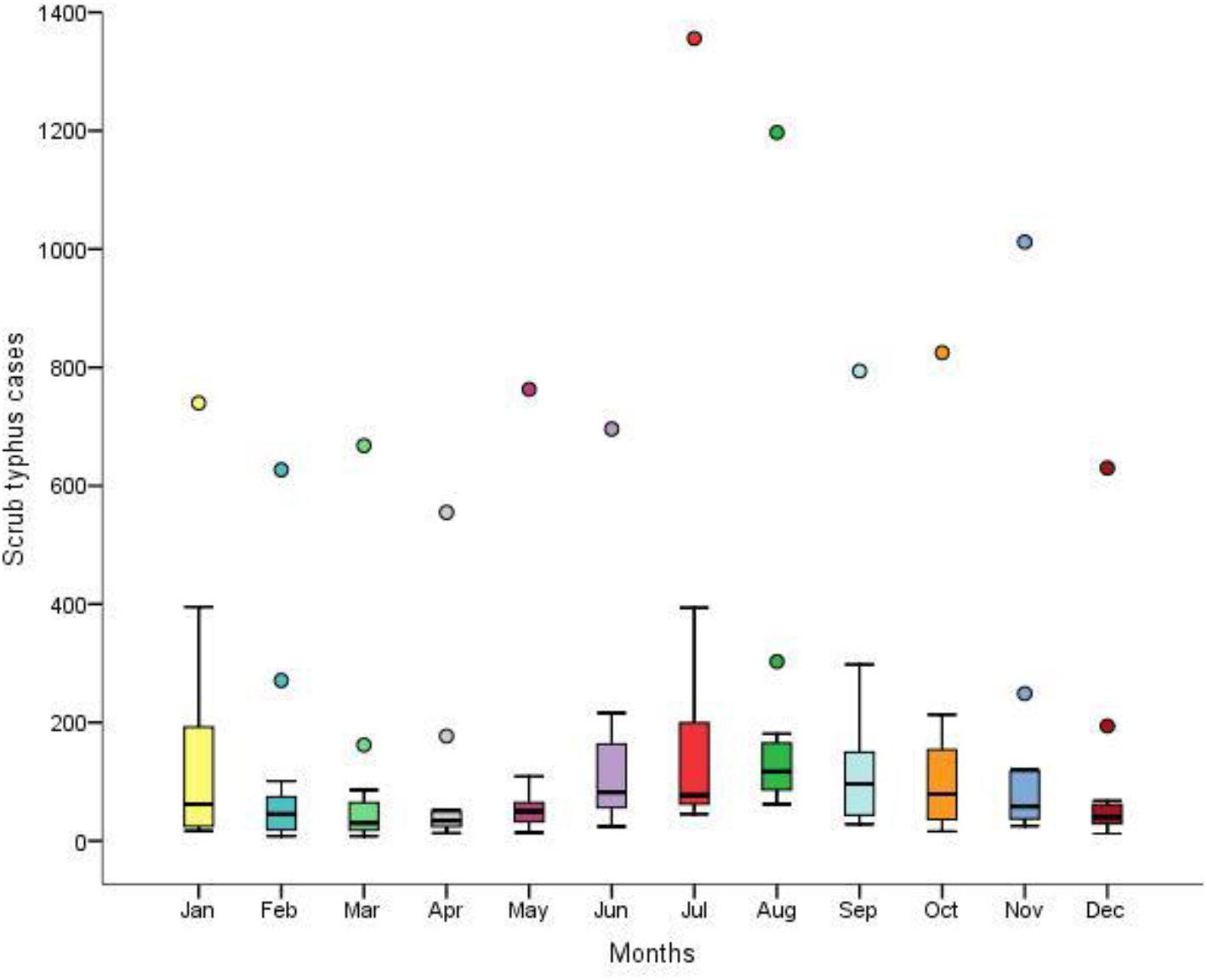
Monthly distribution of scrub typhus cases across the 11 districts of Mizoram (2018-2022). The rectangular box represents the interquartile range; bottom line represents first quartile (25^th^ percentile) and the top line represents third quartile (75^th^ percentile). Median is represented by the horizontal line within the box. Extending from the box are the whiskers (vertical lines). The bottom whisker end represents minimum number of cases and top whisker end represents maximum number of cases which isn’t an outlier. Outliers (mild and extreme) are represented by dots.

### Socio-demographic, occupational and clinical profile of rickettsial infections across the different districts of Mizoram

All age groups (from less than one year to greater than 91 years) were affected; the highly affected group being 31 to 50 years (34.42%), followed by 11 to 30 years (24.33%); the youngest was one month and oldest was 98 years (Supplementary Table 5, Supplementary Figure 1). The proportion of reported cases was slightly higher in males (51.89%) when compared to females (48.11%) (Table 1, Supplementary Table 5). Males were higher than females in scrub typhus and mixed infections, while female cases were more in other rickettsial infections (Table 1, Supplementary Table 5). When stratified based on occupation, 50.73% and 21.74% of the affected individuals were farmers and students, respectively (Supplementary Table 6). Individuals in the fields of business, construction work, and government service make up 21.9% of the reported cases (Supplementary Table 6).

The most common clinical presentations were fever/persistent fever (85.03%), headache (29.35%), rash (26.80%), chills (21.36%), cough (9.21%), body ache (8.58%), and nausea (3.23%) (Supplementary Table 7). Clinical presentations pertaining to CNS (central nervous system) involvement were seen in ∼2% of the cases; the proportion was higher in OX19 positive cases (Supplementary Table 7). Eschar, the pathognomonic lesion, was found in 443 scrub typhus cases, 92 other rickettsial infections, and 46 mixed infection cases (Table 2). The presence of eschar is significantly correlated with other rickettsial infections (p< 0.001), especially in the OX2-positive cases (Table 1 and Supplementary Table 7). While just ∼2% of the reported scrub typhus cases had eschar as a clinical presentation, >7% of the OX2 reactive cases reported eschar (Supplementary Table 7). Despite the nature of the occupation, the distribution of clinical presentations among the different classes was the same (Supplementary Table 8).

Among the reported scrub typhus cases, 19.5% of individuals were hospitalized; the distribution was same across males and females (Supplementary Table 9). Individuals in the age group 31-50 years contributed to >30% of the hospitalized cases (Supplementary Table 9). Even though farmers were high among those who sought care, only <20% of them were hospitalized. On the other hand, the highest proportion of hospitalization (62.62%) was observed in working individuals aged 71 and above (Supplementary Table 9). Table 3 describes the odds of death due to rickettsial infections and Supplementary Table 10 details the case fatality rates. Children aged 10 and below have higher odds of deaths due to rickettsial infections (aOR=5.441, p<0.05) (Table 3). Also, in this age group, CFR due to scrub typhus is high with ∼5 deaths per 1000 (0.51), and is next only to the age group 51-70 whose CFR is 0.66 (Supplementary Table 10). The proportion of rickettsial deaths is more in men (62.96%) than in women (37.04%) (Supplementary Table 10). When compared with the Eastern districts of Mizoram, Western and Central districtshave higher odds (aOR=3.26 and aOR = 2.39, respectively, p< 0.05) of rickettsial deaths (Table3). Among the occupational groups, farmers, business men and construction workers have higher odds (aOR >1, p<0.05) of death if infected with rickettsiae (Table 3). The odds of death are 17 times higher (p<0.001) in hospitalized cases; 76.54% of the hospitalized rickettsial cases suffered death (Table 3, Supplementary Table 10). In the study period, 75 scrub typhus-related deaths were reported; 30.67% of them were hospitalized cases (Supplementary Tables 9 and 10). Scrub typhus deaths were significantly associated with eschar (p<0.05) (Supplementary Table 11). Also, in rickettsial cases with eschar, the odds of deaths are 2.5 times higher (p<0.05) (Table 3). During the study period, scrub typhus CFR was 0.38, while it was 0.11 and 0.28, respectively, for other rickettsial and mixed infections (Supplementary Table 10).

**Table 3.** Socio-demographic and clinical characteristics associated with rickettsial deaths in Mizoram (2018-22)

|  |  |  |  |  |  |  |  |
| --- | --- | --- | --- | --- | --- | --- | --- |
| Variables | Unadjusted odds ratios (uOR) |  |  | Adjusted odds ratios (aOR) |  |  |  |
|  |  | Exp (β) | p-value* | Exp (β) | 95% Confidence Interval |  | p-value* |
|  |  |  |  |  | Lower Bound | Upper Bound |  |
| Eschar presence |  |  |  |  |  |  |  |
|  | Yes | 2.542 | 0.044 | 2.597 | 1.023 | 6.595 | 0.045 |
|  | No (Ref) | 1 |  | 1 |  |  |  |
| Hospitalisation |  |  |  |  |  |  |  |
|  | Yes | 14.821 | 0.000 | 16.995 | 10.025 | 28.811 | 0.000 |
|  | No (Ref) | 1 |  | 1 |  |  |  |
| Age (in years) |  |  |  |  |  |  |  |
|  | Less than 11 | 2.08 | 0.052 | 5.44 | 1.69 | 17.56 | 0.005 |
|  | 11 – 30 (Ref) | 1 |  | 1 |  |  |  |
|  | 31 - 50 | 0.91 | 0.788 | 0.50 | 0.23 | 1.11 | 0.089 |
|  | 51 - 70 | 2.31 | 0.010 | 1.26 | 0.60 | 2.66 | 0.538 |
|  | 71 and more | 1.27 | 0.646 | 0.68 | 0.20 | 2.26 | 0.528 |
| Occupation |  |  |  |  |  |  |  |
|  | Farmer | 1.072 | 0.827 | 3.984 | 1.179 | 13.464 | 0.026 |
|  | Business | 1.957 | 0.089 | 6.693 | 1.898 | 23.605 | 0.003 |
|  | Construction workers | 3.627 | 0.001 | 17.949 | 5.081 | 63.407 | 0.000 |
|  | Government services | .458 | 0.303 | 1.621 | 0.274 | 9.579 | 0.594 |
|  | Pre school | 1.320 | 0.625 | 0.596 | 0.185 | 1.920 | 0.386 |
|  | Others (aged 71 and above) | 1.731 | 0.597 | 2.636 | 0.214 | 32.478 | 0.449 |
|  | Student (Ref) | 1 |  | 1 |  |  |  |
| Sex |  |  |  |  |  |  |  |
|  | Male | 1.579 | 0.047 | 1.559 | 0.981 | 2.480 | 0.060 |
|  | Female (Ref) | 1 |  | 1 |  |  |  |
| Districts |  |  |  |  |  |  |  |
|  | West | 2.575 | 0.044 | 3.265 | 1.287 | 8.281 | 0.013 |
|  | Central | 2.823 | 0.016 | 2.395 | 1.018 | 5.637 | 0.045 |
|  | East (Ref) | 1 |  | 1 |  |  |  |

\*p<0.05 is significant

## Discussion

Despite its importance and public health threat, scrub typhus has stayed under the radar in many parts of India. Studies in the last decade have reported 18781 confirmed cases of scrub typhus across India; most of the studies are hospital-based (138), and only two are community-based [19]. As it is an acute febrile disease that could be easily treated by commonly used antibiotics, many a time the disease goes undiagnosed. The decadal count of 18781 cases might just be a fraction of the true prevalence of scrub typhus across India [19]. The abundance of forest cover, rodents’ easy access to households, farming activities, and conducive environmental conditions provide the ideal setting for the transmission of rickettsial infections in Mizoram [21]. In Mizoram, the first cases of scrub typhus were officially recorded in 2012 [29]. From 2018 onwards, there was a substantial increase in scrub typhus cases. In this study, the Mizoram Government’s initiative to systematically diagnose and record the incidence of scrub typhus and other rickettsial infections across all the testing units has shed important insights on the magnitude of the disease burden in Mizoram; spatial analysis indicates scrub typhus cases are widespread in both urban and rural settings.

Even though there are sporadic reports of scrub typhus across India, there were no systematic recording and reporting at the state level. To our knowledge, this is the first report that details the burden of scrub typhus and other rickettsial infections across an entire state in India. The low rickettsial death rates in Mizoram could be attributed to public awareness and the set-up in place for rapid diagnosis and treatment—key initiatives of the concerned health departments. In India, in the last decade, the case-fatality rate (CFR) of scrub typhus was 6.3%, and in those with multi-organ dysfunction syndrome, the CFR was 38.9% [19]. Delayed presentation, diagnosis and treatment might be the reasons for the high CFR seen in other settings. With the availability of rapid immunochromatographic diagnostic tests [30, 31] (RDT), screening of acute undifferentiated febrile illness (AUFI) for scrub typhus in scrub typhus endemic regions might be rewarding. The sensitivity and specificity of RDT is comparable to ELISA in detecting scrub typhus-specific IgM antibodies [32]. Studies have shown 25.3% of AUFI cases to be due to scrub typhus [19]. As carried out for dengue [33] and chikungunya [34], a nation-wide serosurvey for scrub typhus could select the endemic regions which could be prioritised for integrating scrub typhus screening at district level primary health centers. However, as the rapid diagnostic tests are specific for scrub typhus, they will not detect other rickettsial infections. For this reason, in 2019, Mizoram Govt. introduced WF test across all its testing centers, as patients were reporting with scrub typhus-like symptoms, but were negative when tested with the RDT. Even though WF test has poor sensitivity, it can detect recent or previous infection with typhus group (TG), scrub typhus group (STG) and spotted fever group (SFG) [23]. WF screening has identified a substantial number of other rickettsial and mixed infections in Mizoram. This is in line with serosurveillance data from rodents collected across Mizoram; rodents were seropositive for OX2, OXK, and OX19 antigens [21]. Again, we believe this is the first state-wide systematic recording of other rickettsial infections in India. There were no differences in the clinical presentations between OXK-positive scrub typhus cases and OX2 and/or OX19-positive other rickettsial infections. This indistinguishability calls for increased awareness and necessitation of rickettsia-specific serosurveys to reduce misdiagnosis and understand the true prevalence in the community. Eschar, though described as the “cutaneous hallmark” [35] is not always present in all the infected individuals [3]; thus lowering the suspicion index (a key to early diagnosis) [14, 36]. The prevalence of eschar ranges from 7-80% [8] and is low in South East Asia and India [6]. Dark skin, strain characteristics, and atypical appearances in damp and most skin areas affect the detection and prevalence of eschar [8]. In this study, only 2.5% of the rickettsial cases had eschar, underscoring the unreliability of eschar as a clinical diagnostic marker. The significant association between eschar and death indicates the importance of genome sequencing to correlate strains with clinical severity.

The widespread distribution of rickettsial infections and their continuous antibiotic treatment poses a serious threat of drug resistance in Mizoram. However, because of integrating the screening of rickettsial diseases as part of routine diagnostics across the state, the clinicians might be better placed to detect any deviation in the treatment and recovery period against a specific antibiotic and could course correct. Profiling the genome of the circulating strains and correlating it with disease severity are critical in devising prevention, control and treatment strategies. As screening of scrub typhus is not routinely carried out in many parts of the country, indiscriminate antibiotic use against AUFI might lead to emergence of drug resistant rickettsiae.

In this study, the majority of the rickettsial infections were reported in monsoon and post-monsoon seasons, and is in line with several studies carried out in India [37-39]. Similarly, as reported earlier, the majority of the cases were in individuals involved in farming activities [39-41]. In Mizoram, Jhum or shifting cultivation, also known as slash and burn agriculture is the major farming activity, and an estimated 54% are practicing it [42]. Even though 50.73% of the cases are farmers, only 16.33% of them were hospitalized. The reason for low hospitalization among farmers might be partial immunity due to repeated exposure to natural infection in the fields. Natural *Orientia* infection does not elicit sterilizing and long lasting immunity, and does not confer cross protection to infection from other strains [43]. High hospitalization (62.62%) in adults >71 years may be because of their lack of prior exposure (the first scrub typhus cases were recorded only in 2012), and age-related immunosenescence [44].

Due to the resource-limited settings, the study has several limitations. Due to the poor sensitivity of the WF test, the true burden of rickettsial infections might be substantially high. One alternative is to use the highly sensitive rapid immunochromatographic test for scrub typhus. If negative, the samples could be tested by ELISA specific to SFG and TG rickettsiae; this approach would be cost-effective, and increase the sensitivity and specificity of detection. Also, the decline in caseloads in 2020 and 2021 could be due to the pandemic as all testing centers were engaged with the diagnosis of Covid-19.

Overall, this study details the importance of integrating the screening of rickettsial diseases at district level across all the health centers. This approach could serve as a template for other states/regions which are endemic to scrub typhus and other rickettsial infections.

## Data Availability

All data produced in the present study are available upon reasonable request to the authors

## Ethics statement

The ethical approval to use the data was provided by Directorate of Health Services, Government of Mizoram (NO. D.32020/412014-DHS/IDSP).

## Acknowledgement

The authors would like to convey their sincere gratitude to health officials and other concerned Govt. officials of Mizoram for making provision to distribute WF and ICT kits to diagnose scrub typhus and other rickettsial diseases to all Govt. hospitals in the state free of cost. The authors also sincerely thank the Director of Health Services, Govt. of Mizoram, for giving permission to collect and analyze the secondary data. The team would also like to convey their sincere thanks to all district health officials, data managers, and data entry operators under IDSP for their continuous and untiring support in collection, entry, and reporting of the data. The authors also acknowledge the Institutional Advanced-level Biotech Hub (BT/NER/143/SP44393/2021, dated 18^th^ November 2022) Dept. of Zoology, Pachhunga University College, for providing computational and laboratory facilities, Science and Engineering Research Board, New Delhi, for providing financial support under Core Research Grant (CRG/2019/003016/BHS; dated 7^th^ February 2022) and DHR (R.11013/11/2021-GIA/HR) for funding.

## Author contributions

V, LP and PBN conceptualized, designed, and wrote the first draft; KB performed the GIS analysis; CA and NKK performed statistics; LP, GR, DKS, H and YM contributed to the literature search, review and editing of the manuscript; All authors have read and agreed to the final version of the manuscript.

## References

1. Scrub Typhus [https://www.cdc.gov/typhus/scrub/index.html#:~:text=Scrub%20typhus%2C%20also%20known%20as,body%20aches%2C%20and%20sometimes%20rash.]

2. Rajapakse S, Weeratunga P, Sivayoganathan S, Fernando SD: Clinical manifestations of scrub typhus. Transactions of the Royal Society of Tropical Medicine and Hygiene 2017, 111(2):43–54.

3. Taylor AJ, Paris DH, Newton PN: A Systematic Review of Mortality from Untreated Scrub Typhus (Orientia tsutsugamushi). PLoS neglected tropical diseases 2015, 9(8):e0003971.

4. Mahajan SK: Scrub typhus. The Journal of the Association of Physicians of India 2005, 53:954–958.

5. John R, Varghese GM: Scrub typhus: a reemerging infection. Curr Opin Infect Dis 2020, 33(5):365–371.

6. Venkategowda PM, Rao SM, Mutkule DP, Rao MV, Taggu AN: Scrub typhus: Clinical spectrum and outcome. Indian J Crit Care Med 2015, 19(4):208–213.

7. Saini R, Pui JC, Burgin S: Rickettsialpox: report of three cases and a review. J Am Acad Dermatol 2004, 51(5 Suppl):S137–142.

8. Rajapakse S, Rodrigo C, Fernando D: Scrub typhus: pathophysiology, clinical manifestations and prognosis. Asian Pac J Trop Med 2012, 5(4):261–264.

9. Vivekanandan M, Mani A, Priya YS, Singh AP, Jayakumar S, Purty S: Outbreak of scrub typhus in Pondicherry. The Journal of the Association of Physicians of India 2010, 58:24–28.

10. Kim DM, Kim SW, Choi SH, Yun NR: Clinical and laboratory findings associated with severe scrub typhus. BMC infectious diseases 2010, 10:108.

11. Kim D-M: Clinical Features and Diagnosis of Scrub Typhus. Infection and Chemotherapy 2009, 41(6):315–322.

12. Stephen S, Ambroise S, Gunasekaran D, Hanifah M, Sangeetha B, Pradeep J, Sarangapani K: Serological evidence of spotted fever group rickettsiosis in and around Puducherry, south India—A three years study. Journal of vector borne diseases 2018, 55(2):144–150.

13. Mane A, Kamble S, Singh MK, Ratnaparakhi M, Nirmalkar A, Gangakhedkar R: Seroprevalence of spotted fever group and typhus group rickettsiae in individuals with acute febrile illness from Gorakhpur, India. International Journal of Infectious Diseases 2019, 79:195–198.

14. Rathi N, Rathi A: Rickettsial Infections: Indian Perspective. Indian pediatrics 2010(47):157–155.

15. Snowden J, Ladd M, King KC: Rickettsial Infection. In: StatPearls. Treasure Island (FL); 2023.

16. Lalrinkima H, Lalremruata R, Lalchhandama C, Khiangte L, Siamthara FH, Lalnunpuia C, Borthakur SK, Patra G: Scrub typhus in Mizoram, India. Journal of vector borne diseases 2017, 54(4):369–371.

17. MIZORAM STATE BIODIVERSITY BOARD [https://forest.mizoram.gov.in/page/mizoram-state-biodiversity-board]

18. >Biological Diversity In Mizoram [http://mizenvis.nic.in/Database/Biodiversity_1444.aspx]

19. Devasagayam E, Dayanand D, Kundu D, Kamath MS, Kirubakaran R, Varghese GM: The burden of scrub typhus in India: A systematic review. PLoS neglected tropical diseases 2021, 15(7):e0009619.

20. Mizoram [http://slbcne.nic.in/mizoram/MIZORAMstateprofile.pdf]

21. Pautu L, Lalmalsawma P, Vanramliana, Balasubramani K, Balabaskaran Nina P, Rosangkima G, Sarma DK, Malvi Y, Hunropuia: Seroprevalence of scrub typhus and other rickettsial diseases among the household rodents of Mizoram, North-East India. Zoonoses Public Health 2023.

22. Anitharaj V, Stephen S, Pradeep J, Park S, Kim SH, Kim YJ, Kim EY, Kim YW: Serological Diagnosis of Acute Scrub Typhus in Southern India: Evaluation of InBios Scrub Typhus Detect IgM Rapid Test and Comparison with other Serological Tests. J Clin Diagn Res 2016, 10(11):DC07–DC10.

23. Cox AL, Tadi P: Weil Felix Test. In: StatPearls. Treasure Island (FL); 2022.

24. Mittal V, Gupta N, Bhattacharya D, Kumar K, Ichhpujani RL, Singh S, Chhabra M, Rana UV: Serological evidence of rickettsial infections in Delhi. The Indian journal of medical research 2012, 135(4):538–541.

25. Tenny S, Boktor SW: Incidence. In: StatPearls. Treasure Island (FL); 2023.

26. Wang W, Kim R, Subramanian SV: Population Estimates for Districts and Parliamentary Constituencies in India, 2020. In., V1 edn: Harvard Dataverse; 2021.

27. Lalmalsawma P, Balasubramani K, James MM, Pautu L, Prasad KA, Sarma DK, Balabaskaran Nina P: Malaria hotspots and climate change trends in the hyper-endemic malaria settings of Mizoram along the India-Bangladesh borders. Scientific reports 2023, 13(1):4538.

28. How Hot Spot Analysis (Getis-Ord Gi*) works [https://pro.arcgis.com/en/pro-app/latest/tool-reference/spatial-statistics/h-how-hot-spot-analysis-getis-ord-gi-spatial-stati.htm]

29. Lalmalsawma P, Pautu L: Scenario of Scrub typhus disease in Mizoram, Northeast India. International Journal of Current Advanced Research 2017, 6:6341–6344.

30. Kim YJ, Park S, Premaratna R, Selvaraj S, Park SJ, Kim S, Kim D, Kim MS, Shin DH, Choi KC et al: Clinical Evaluation of Rapid Diagnostic Test Kit for Scrub Typhus with Improved Performance. J Korean Med Sci 2016, 31(8):1190–1196.

31. Silpasakorn S, Waywa D, Hoontrakul S, Suttinont C, Losuwanaluk K, Suputtamongkol Y: Performance of SD Bioline Tsutsugamushi assays for the diagnosis of scrub typhus in Thailand. J Med Assoc Thai 2012, 95 Suppl 2:S18–22.

32. Kannan K, John R, Kundu D, Dayanand D, Abhilash KPP, Mathuram AJ, Zachariah A, Sathyendra S, Hansdak SG, Abraham OC et al: Performance of molecular and serologic tests for the diagnosis of scrub typhus. PLoS neglected tropical diseases 2020, 14(11):e0008747.

33. Murhekar MV, Kamaraj P, Kumar MS, Khan SA, Allam RR, Barde P, Dwibedi B, Kanungo S, Mohan U, Mohanty SS et al: Burden of dengue infection in India, 2017: a cross-sectional population based serosurvey. The Lancet Global health 2019, 7(8):e1065–e1073.

34. Kumar MS, Kamaraj P, Khan SA, Allam RR, Barde PV, Dwibedi B, Kanungo S, Mohan U, Mohanty SS, Roy S et al: Seroprevalence of chikungunya virus infection in India, 2017: a cross-sectional population-based serosurvey. Lancet Microbe 2021, 2(1):e41–e47.

35. Basu S, Chakravarty A: Neurological Manifestations of Scrub Typhus. Curr Neurol Neurosci Rep 2022, 22(8):491–498.

36. Khan SA, Murhekar MV, Bora T, Kumar S, Saikia J, Kamaraj P, Sabarinanthan R: Seroprevalence of Rickettsial Infections in Northeast India: A Population-Based Cross-Sectional Survey. Asia Pacific Journal of Public Health 2021, 33(5):516–522.

37. Murhekar MV, Vivian Thangaraj JW, Sadanandane C, Mittal M, Gupta N, Rose W, Sahay S, Kant R, Gupte MD: Investigations of seasonal outbreaks of acute encephalitis syndrome due to Orientia tsutsugamushi in Gorakhpur region, India: A One Health case study. The Indian journal of medical research 2021, 153(3):375–381.

38. Nallasamy K, Gupta S, Bansal A, Biswal M, Jayashree M, Zaman K, Williams V, Kumar A: Clinical Profile and Predictors of Intensive Care Unit Admission in Pediatric Scrub Typhus: A Retrospective Observational Study from North India. Indian J Crit Care Med 2020, 24(6):445–450.

39. Narang R, Deshmukh P, Jain J, Jain M, Raut A, Deotale V, Pote K, Rahi M: Scrub typhus in urban areas of Wardha district in central India. The Indian journal of medical research 2022, 156(3):435–441.

40. Saha B, Chatterji S, Mitra K, Ghosh S, Naskar A, Ghosh MK, Parui S, Thakur A, Bhattacharya B, Majumdar D et al: Socio-demographic and Clinico-Epidemiological Study of Scrub Typhus in Two Tertiary Care Hospitals of Kolkata. The Journal of the Association of Physicians of India 2018, 66(5):22–25.

41. Jakharia A, Borkakoty B, Biswas D, Yadav K, Mahanta J: Seroprevalence of Scrub Typhus Infection in Arunachal Pradesh, India. Vector borne and zoonotic diseases 2016, 16(10):659–663.

42. Sati VP: Shifting cultivation in Mizoram, India: An empirical study of its economic implications. Journal of Mountain Science 2019, 16(9):2136–2149.

43. Paris DH, Shelite TR, Day NP, Walker DH: Unresolved problems related to scrub typhus: a seriously neglected life-threatening disease. The American journal of tropical medicine and hygiene 2013, 89(2):301–307.

44. Weyand CM, Goronzy JJ: Aging of the Immune System. Mechanisms and Therapeutic Targets. Annals of the American Thoracic Society 2016, 13 Suppl 5(Suppl 5):S422–S428.

