## Supplementary Table 1 for "Epidemiology of scrub typhus and other rickettsial infections (2018-22) in the hyper-endemic setting of Mizoram, North-East India"

**Supplementary Table 1 Monthly distribution of scrub typhus cases across the districts of Mizoram (2018-2022)**

|  | 2018 | | | | | | | | | | | | | |
| --- | --- | --- | --- | --- | --- | --- | --- | --- | --- | --- | --- | --- | --- | --- |
|  | **Jan** | **Feb** | **March** | **April** | **May** | **June** | **July** | **Aug** | **Sep** | **Oct** | **Nov** | **Dec** | **Total** | **(%)** |
| Aizawl | 116 | 155 | 113 | 41 | 22 | 30 | 247 | 240 | 38 | 1 | 6 | 1 | 1010 | 46.27 |
| Champhai | 0 | 0 | 1 | 0 | 0 | 0 | 12 | 13 | 8 | 0 | 0 | 0 | 34 | 1.56 |
| Hnahthial | 5 | 0 | 0 | 0 | 0 | 22 | 0 | 0 | 0 | 0 | 0 | 0 | 27 | 1.24 |
| Khawzawl | 1 | 0 | 0 | 0 | 0 | 0 | 4 | 1 | 3 | 0 | 0 | 0 | 9 | 0.41 |
| Kolasib | 1 | 1 | 0 | 0 | 0 | 3 | 11 | 7 | 0 | 0 | 0 | 0 | 23 | 1.05 |
| Lawngtlai | 12 | 2 | 0 | 0 | 3 | 10 | 17 | 6 | 11 | 30 | 21 | 7 | 119 | 5.45 |
| Lunglei | 190 | 0 | 0 | 2 | 7 | 87 | 6 | 0 | 0 | 8 | 0 | 0 | 300 | 13.74 |
| Mamit | 3 | 19 | 3 | 2 | 16 | 10 | 13 | 9 | 1 | 0 | 2 | 0 | 78 | 3.57 |
| Saitual | 28 | 21 | 0 | 0 | 0 | 0 | 5 | 4 | 2 | 0 | 2 | 0 | 62 | 2.84 |
| Serchhip | 2 | 2 | 4 | 3 | 5 | 31 | 184 | 74 | 59 | 15 | 27 | 107 | 513 | 23.50 |
| Siaha | 6 | 0 | 2 | 0 | 0 | 0 | 0 | 0 | 0 | 0 | 0 | 0 | 8 | 0.37 |
|  | 364 | 200 | 123 | 48 | 53 | 193 | 499 | 354 | 122 | 54 | 58 | 115 | **2183** |  |
| (%) | 16.67 | 9.16 | 5.63 | 2.20 | 2.43 | 8.84 | 22.86 | 16.22 | 5.59 | 2.47 | 2.66 | 5.27 |  |  |
|  | **2019** | | | | | | | | | | | | | |
|  | **Jan** | **Feb** | **March** | **April** | **May** | **June** | **July** | **Aug** | **Sep** | **Oct** | **Nov** | **Dec** | **Total** | **(%)** |
| Aizawl | 254 | 151 | 223 | 196 | 205 | 259 | 523 | 478 | 421 | 398 | 484 | 287 | 3879 | 66.21 |
| Champhai | 1 | 0 | 1 | 2 | 1 | 2 | 0 | 8 | 5 | 8 | 3 | 1 | 32 | 0.55 |
| Hnahthial | 0 | 0 | 0 | 0 | 0 | 0 | 0 | 0 | 2 | 0 | 0 | 1 | 3 | 0.05 |
| Khawzawl | 14 | 0 | 0 | 0 | 0 | 0 | 0 | 4 | 1 | 1 | 1 | 0 | 21 | 0.36 |
| Kolasib | 0 | 0 | 0 | 0 | 0 | 0 | 3 | 44 | 5 | 3 | 0 | 0 | 55 | 0.94 |
| Lawngtlai | 10 | 10 | 2 | 5 | 0 | 2 | 49 | 17 | 23 | 6 | 1 | 0 | 125 | 2.13 |
| Lunglei | 1 | 5 | 2 | 1 | 4 | 1 | 2 | 4 | 7 | 14 | 14 | 4 | 59 | 1.01 |
| Mamit | 8 | 4 | 6 | 0 | 3 | 2 | 5 | 8 | 22 | 39 | 9 | 0 | 106 | 1.81 |
| Saitual | 4 | 5 | 3 | 8 | 14 | 23 | 33 | 65 | 65 | 22 | 13 | 4 | 259 | 4.42 |
| Serchhip | 235 | 186 | 126 | 136 | 38 | 84 | 48 | 36 | 105 | 100 | 132 | 39 | 1265 | 21.59 |
| Siaha | 3 | 0 | 1 | 1 | 1 | 3 | 3 | 7 | 5 | 4 | 15 | 12 | 55 | 0.94 |
|  | 530 | 361 | 364 | 349 | 266 | 376 | 666 | 671 | 661 | 595 | 672 | 348 | **5859** |  |
| (%) | 9.05 | 6.16 | 6.21 | 5.96 | 4.54 | 6.42 | 11.37 | 11.45 | 11.28 | 10.16 | 11.47 | 5.94 |  |  |
| 2020 | | | | | | | | | | | | | | |
|  | **Jan** | **Feb** | **March** | **April** | **May** | **June** | **July** | **Aug** | **Sep** | **Oct** | **Nov** | **Dec** | **Total** | **(%)** |
| Aizawl | 121 | 104 | 70 | 46 | 83 | 157 | 214 | 160 | 148 | 224 | 249 | 172 | 1748 | 63.70 |
| Champhai | 4 | 1 | 3 | 2 | 0 | 2 | 2 | 3 | 1 | 5 | 7 | 7 | 37 | 1.35 |
| Hnahthial | 0 | 0 | 0 | 0 | 0 | 0 | 0 | 3 | 0 | 0 | 1 | 0 | 4 | 0.15 |
| Khawzawl | 0 | 0 | 1 | 0 | 2 | 0 | 6 | 11 | 4 | 6 | 16 | 1 | 47 | 1.71 |
| Kolasib | 2 | 0 | 0 | 0 | 0 | 3 | 1 | 2 | 3 | 1 | 2 | 0 | 14 | 0.51 |
| Lawngtlai | 30 | 20 | 11 | 5 | 23 | 16 | 66 | 21 | 31 | 36 | 24 | 3 | 286 | 10.42 |
| Lunglei | 5 | 3 | 1 | 2 | 6 | 18 | 26 | 4 | 14 | 23 | 22 | 12 | 136 | 4.96 |
| Mamit | 2 | 0 | 0 | 0 | 4 | 4 | 10 | 33 | 48 | 46 | 35 | 25 | 207 | 7.54 |
| Saitual | 12 | 3 | 1 | 1 | 4 | 6 | 13 | 13 | 2 | 14 | 10 | 0 | 79 | 2.88 |
| Serchhip | 104 | 30 | 0 | 0 | 0 | 0 | 0 | 0 | 0 | 0 | 0 | 0 | 134 | 4.88 |
| Siaha | 5 | 4 | 1 | 3 | 0 | 5 | 4 | 3 | 7 | 9 | 5 | 6 | 52 | 1.90 |
|  | 285 | 165 | 88 | 59 | 122 | 211 | 342 | 253 | 258 | 364 | 371 | 226 | **2744** |  |
| (%) | 10.39 | 6.01 | 3.21 | 2.15 | 4.45 | 7.69 | 12.46 | 9.22 | 9.40 | 13.27 | 13.52 | 8.24 |  |  |
| 2021 | | | | | | | | | | | | | | |
|  | **Jan** | **Feb** | **March** | **April** | **May** | **June** | **July** | **Aug** | **Sep** | **Oct** | **Nov** | **Dec** | **Total** | **(%)** |
| Aizawl | 114 | 112 | 129 | 62 | 81 | 121 | 145 | 116 | 89 | 80 | 160 | 83 | 1292 | 55.62 |
| Champhai | 6 | 8 | 5 | 2 | 4 | 0 | 0 | 2 | 3 | 5 | 8 | 8 | 51 | 2.20 |
| Hnahthial | 0 | 0 | 1 | 1 | 2 | 0 | 2 | 2 | 2 | 4 | 2 | 1 | 17 | 0.73 |
| Khawzawl | 3 | 3 | 4 | 0 | 1 | 0 | 0 | 0 | 0 | 0 | 0 | 0 | 11 | 0.47 |
| Kolasib | 4 | 0 | 2 | 3 | 1 | 1 | 1 | 11 | 4 | 0 | 6 | 1 | 34 | 1.46 |
| Lawngtlai | 35 | 32 | 28 | 18 | 15 | 10 | 13 | 8 | 1 | 4 | 8 | 15 | 187 | 8.05 |
| Lunglei | 14 | 5 | 10 | 13 | 18 | 23 | 22 | 7 | 15 | 17 | 20 | 13 | 177 | 7.62 |
| Mamit | 12 | 10 | 2 | 2 | 5 | 5 | 6 | 9 | 9 | 23 | 15 | 2 | 100 | 4.30 |
| Saitual | 2 | 0 | 0 | 1 | 0 | 0 | 1 | 9 | 3 | 0 | 4 | 2 | 22 | 0.95 |
| Serchhip | 32 | 28 | 18 | 15 | 4 | 19 | 32 | 59 | 35 | 27 | 37 | 22 | 328 | 14.12 |
| Siaha | 6 | 0 | 1 | 7 | 7 | 1 | 10 | 11 | 10 | 15 | 15 | 21 | 104 | 4.48 |
|  | 228 | 198 | 200 | 124 | 138 | 180 | 232 | 234 | 171 | 175 | 275 | 168 | **2323** |  |
| (%) | 9.81 | 8.52 | 8.61 | 5.34 | 5.94 | 7.75 | 9.99 | 10.07 | 7.36 | 7.53 | 11.84 | 7.23 |  |  |
| 2022 | | | | | | | | | | | | | | |
|  | **Jan** | **Feb** | **March** | **April** | **May** | **June** | **July** | **Aug** | **Sep** | **Oct** | **Nov** | **Dec** | **Total** | **(%)** |
| Aizawl | 135 | 105 | 133 | 210 | 372 | 129 | 227 | 203 | 98 | 122 | 113 | 87 | 1934 | 29.56 |
| Champhai | 6 | 4 | 1 | 7 | 16 | 20 | 31 | 66 | 32 | 34 | 20 | 16 | 253 | 3.87 |
| Hnahthial | 12 | 15 | 16 | 19 | 12 | 40 | 75 | 112 | 92 | 33 | 30 | 24 | 480 | 7.34 |
| Khawzawl | 2 | 5 | 3 | 34 | 29 | 83 | 48 | 65 | 30 | 28 | 19 | 19 | 365 | 5.58 |
| Kolasib | 55 | 21 | 17 | 42 | 32 | 45 | 33 | 32 | 16 | 12 | 17 | 11 | 333 | 5.09 |
| Lawngtlai | 55 | 37 | 45 | 18 | 24 | 119 | 124 | 129 | 38 | 20 | 22 | 9 | 640 | 9.78 |
| Lunglei | 32 | 32 | 30 | 34 | 9 | 41 | 65 | 47 | 39 | 39 | 59 | 26 | 453 | 6.92 |
| Mamit | 33 | 15 | 27 | 19 | 23 | 40 | 41 | 83 | 105 | 105 | 59 | 41 | 591 | 9.03 |
| Saitual | 25 | 19 | 17 | 15 | 46 | 53 | 78 | 58 | 42 | 43 | 29 | 47 | 472 | 7.21 |
| Serchhip | 22 | 25 | 14 | 23 | 62 | 82 | 130 | 134 | 99 | 65 | 53 | 26 | 735 | 11.24 |
| Siaha | 9 | 30 | 25 | 15 | 42 | 41 | 50 | 53 | 11 | 4 | 5 | 1 | 286 | 4.37 |
|  | 386 | 308 | 328 | 436 | 667 | 693 | 902 | 982 | 602 | 505 | 426 | 307 | **6542** |  |
| (%) | 5.90 | 4.71 | 5.01 | 6.66 | 10.20 | 10.59 | 13.79 | 15.01 | 9.20 | 7.72 | 6.51 | 4.69 |  |  |
