## Supplementary Table 2 for "Epidemiology of scrub typhus and other rickettsial infections (2018-22) in the hyper-endemic setting of Mizoram, North-East India"

**Supplementary Table 2 Weil Felix test based diagnosis of rickettsial infections across the districts of Mizoram (2019-2022)**

|  | Scrub typhus | | Other rickettsial infections | | | | Mixed (scrub typhus and other rickettsial) infections | | |  |
| --- | --- | --- | --- | --- | --- | --- | --- | --- | --- | --- |
| Districts | **OXK** | | **OX2** | **OX19** | **OX19 & OX2** | | **OXK & OX19** | **OXK & OX2** | **OXK, OX2& OX19** | **Total** |
|  | **5838** | | **1068** | **572** | **206** | | **331** | **466** | **620** | **9101** |
| 2019 | | | | | | | | | | |
| Aizawl | 150 | | 6 | 5 | | 0 | 0 | 2 | 82 | 245 |
| Champhai | 17 | | 44 | 19 | | 3 | 1 | 0 | 0 | 84 |
| Hnahthial | 3 | | 104 | 4 | | 3 | 0 | 11 | 2 | 127 |
| Khawzawl | 11 | | 97 | 135 | | 21 | 1 | 0 | 0 | 265 |
| Kolasib | 55 | | 10 | 5 | | 12 | 5 | 6 | 4 | 97 |
| Lawngtlai | 0 | | 0 | 0 | | 0 | 0 | 15 | 0 | 15 |
| Lunglei | 0 | | 0 | 0 | | 0 | 0 | 0 | 0 | 0 |
| Mamit | 0 | | 0 | 0 | | 0 | 0 | 0 | 12 | 12 |
| Saitual | 0 | | 0 | 0 | | 0 | 0 | 0 | 0 | 0 |
| Serchhip | 61 | | 0 | 1 | | 0 | 0 | 0 | 0 | 62 |
| Siaha | 0 | | 2 | 1 | | 0 | 0 | 0 | 0 | 3 |
|  | **297** | | **263** | **170** | | **39** | **7** | **34** | **100** | **910** |
|  | **2020** | | | | | | | | | |
|  | **OXK** | | **OX2** | **OX19** | | **OX19 & OX2** | **OXK & OX19** | **OXK & OX2** | **OXK, OX2& OX19** | **Total** |
| Aizawl | 407 | | 0 | 1 | | 2 | 1 | 17 | 196 | 624 |
| Champhai | 30 | | 35 | 8 | | 0 | 1 | 0 | 2 | 76 |
| Hnahthial | 4 | | 142 | 3 | | 1 | 1 | 5 | 0 | 156 |
| Khawzawl | 47 | | 16 | 110 | | 11 | 8 | 10 | 1 | 203 |
| Kolasib | 12 | | 38 | 7 | | 26 | 4 | 20 | 5 | 112 |
| Lawngtlai | 0 | | 0 | 0 | | 0 | 0 | 48 | 0 | 48 |
| Lunglei | 0 | | 68 | 2 | | 1 | 0 | 0 | 0 | 71 |
| Mamit | 186 | | 86 | 53 | | 8 | 14 | 2 | 12 | 361 |
| Saitual | 38 | | 0 | 0 | | 0 | 0 | 0 | 0 | 38 |
| Serchhip | 10 | | 0 | 0 | | 0 | 0 | 0 | 0 | 10 |
| Siaha | 5 | | 4 | 4 | | 0 | 0 | 0 | 0 | 13 |
|  | **739** | | **389** | **188** | | **49** | **29** | **102** | **216** | **1712** |
|  | **2021** | | | | | | | | | |
|  | **OXK** | | **OX2** | **OX19** | | **OX19 & OX2** | **OXK & OX19** | **OXK & OX2** | **OXK, OX2& OX19** | **Total** |
| Aizawl | 473 | | 79 | 18 | | 6 | 4 | 50 | 99 | 729 |
| Champhai | 37 | | 30 | 6 | | 5 | 7 | 2 | 2 | 89 |
| Hnahthial | 17 | | 40 | 4 | | 2 | 1 | 19 | 0 | 83 |
| Khawzawl | 11 | | 6 | 7 | | 0 | 0 | 2 | 0 | 26 |
| Kolasib | 34 | | 36 | 18 | | 13 | 2 | 24 | 2 | 129 |
| Lawngtlai | 1 | | 0 | 0 | | 0 | 82 | 17 | 0 | 100 |
| Lunglei | 8 | | 57 | 1 | | 0 | 0 | 0 | 0 | 66 |
| Mamit | 66 | | 11 | 19 | | 1 | 22 | 8 | 2 | 129 |
| Saitual | 20 | | 39 | 11 | | 2 | 2 | 16 | 0 | 90 |
| Serchhip | 202 | | 11 | 7 | | 1 | 1 | 2 | 1 | 225 |
| Siaha | 1 | | 0 | 0 | | 0 | 0 | 0 | 0 | 1 |
|  | **870** | | **309** | **91** | | **30** | **121** | **140** | **106** | **1667** |
|  | **2022** | | | | | | | | | |
|  | **OXK** | **OX2** | | **OX19** | | **OX19 & OX2** | **OXK & OX19** | **OXK & OX2** | **OXK, OX2& OX19** | **Total** |
| Aizawl | 913 | 10 | | 11 | | 11 | 29 | 14 | 74 | 1062 |
| Champhai | 108 | 6 | | 6 | | 2 | 7 | 4 | 4 | 137 |
| Hnahthial | 442 | 25 | | 14 | | 8 | 14 | 46 | 9 | 558 |
| Khawzawl | 360 | 19 | | 21 | | 9 | 12 | 22 | 20 | 463 |
| Kolasib | 271 | 6 | | 12 | | 8 | 12 | 12 | 1 | 322 |
| Lawngtlai | 140 | 1 | | 5 | | 4 | 20 | 6 | 8 | 184 |
| Lunglei | 82 | 6 | | 3 | | 5 | 6 | 8 | 5 | 115 |
| Mamit | 565 | 12 | | 30 | | 16 | 35 | 30 | 39 | 727 |
| Saitual | 445 | 7 | | 10 | | 14 | 13 | 20 | 15 | 524 |
| Serchhip | 598 | 15 | | 11 | | 11 | 26 | 28 | 22 | 711 |
| Siaha | 8 | 0 | | 0 | | 0 | 0 | 0 | 1 | 9 |
|  | **3932** | **107** | | **123** | | **88** | **174** | **190** | **198** | **4812** |
