## Supplementary Table 3 for "Epidemiology of scrub typhus and other rickettsial infections (2018-22) in the hyper-endemic setting of Mizoram, North-East India"

**Supplementary Table 3** **Incidence rates of scrub typhus, other rickettsial and mixed infections across the districts of Mizoram (2018-2022)**

| Cases / 1000 persons-year | Aizawl | | Champhai | Hnahthial | Khawzawl | Kolasib | Lawngtlai | Lunglei | Mamit | Saitual | Serchhip | Siaha | Mizoram |
| --- | --- | --- | --- | --- | --- | --- | --- | --- | --- | --- | --- | --- | --- |
| Scrub typhus |  | |  |  |  |  |  |  |  |  |  |  |  |
| 2018 | 2.21 | | 0.33 | 0.76 | 0.24 | 0.21 | 0.80 | 2.17 | 0.80 | 1.08 | 8.05 | 0.17 | 1.69 |
| 2019 | 8.49 | | 0.31 | 0.08 | 0.56 | 0.51 | 0.85 | 0.43 | 1.08 | 4.53 | 19.86 | 1.17 | 4.53 |
| 2020 | 3.82 | | 0.36 | 0.11 | 1.26 | 0.13 | 1.93 | 0.98 | 2.11 | 1.38 | 2.10 | 1.10 | 2.12 |
| 2021 | 2.83 | | 0.49 | 0.48 | 0.30 | 0.31 | 1.26 | 1.28 | 1.02 | 0.38 | 5.15 | 2.20 | 1.79 |
| 2022 | 4.23 | | 2.44 | 13.42 | 9.80 | 3.07 | 4.33 | 3.28 | 6.03 | 8.25 | 11.54 | 6.06 | 5.05 |
| Avg. incidence rate | 4.32 | | 0.79 | 2.97 | 2.43 | 0.85 | 1.84 | 1.63 | 2.21 | 3.12 | 9.34 | 2.14 | 3.04 |
| Other rickettsial infections | |  |  |  |  |  |  |  |  |  |  |  |  |
| 2018 | - | | - | - | - | - | - | - | - | - | - | - | - |
| 2019 | 0.02 | | 0.64 | 3.10 | 6.79 | 0.25 | 0.00 | 0.00 | 0.00 | 0.00 | 0.02 | 0.06 | 0.36 |
| 2020 | 0.01 | | 0.42 | 4.08 | 3.68 | 0.65 | 0.00 | 0.51 | 1.50 | 0.00 | 0.00 | 0.17 | 0.01 |
| 2021 | 0.02 | | 0.04 | 0.13 | 0.03 | 0.06 | 0.00 | 0.04 | 0.03 | 0.09 | 0.03 | 0.00 | 0.03 |
| 2022 | 0.07 | | 0.14 | 1.31 | 1.32 | 0.24 | 0.07 | 0.10 | 0.59 | 0.54 | 0.58 | 0.00 | 0.25 |
| Avg. incidence rate | 0.08 | | 0.40 | 2.45 | 3.03 | 0.44 | 0.02 | 0.26 | 0.60 | 0.36 | 0.22 | 0.06 | 0.36 |
| Mixed (scrub typhus and other rickettsial) infections | | |  |  |  |  |  |  |  |  |  |  |  |
| 2018 | - | | - | - | - | - | - | - | - | - | - | - | - |
| 2019 | 0.18 | | 0.01 | 0.36 | 0.03 | 0.14 | 0.10 | 0.00 | 0.12 | 0.00 | 0.00 | 0.00 | 0.11 |
| 2020 | 0.47 | | 0.03 | 0.17 | 0.51 | 0.27 | 0.32 | 0.00 | 0.29 | 0.00 | 0.00 | 0.00 | 0.27 |
| 2021 | 0.33 | | 0.11 | 0.56 | 0.05 | 0.26 | 0.67 | 0.00 | 0.33 | 0.31 | 0.06 | 0.00 | 0.28 |
| 2022 | 0.26 | | 0.14 | 1.93 | 1.45 | 0.23 | 0.23 | 0.14 | 1.06 | 0.84 | 1.19 | 0.02 | 0.43 |
| Avg. incidence rate | 0.31 | | 0.07 | 0.76 | 0.51 | 0.22 | 0.33 | 0.03 | 0.45 | 0.29 | 0.31 | 0.01 | 0.27 |
