## Supplementary Table 4 for "Epidemiology of scrub typhus and other rickettsial infections (2018-22) in the hyper-endemic setting of Mizoram, North-East India"

**Supplementary Table 4 Incidence rates of rickettsial infections across districts of Mizoram based on antigenic strains (2019-2022)**

| Cases/1000 persons-year | Aizawl | Champhai | Hnahthial | Khawzawl | Kolasib | Lawngtlai | Lunglei | Mamit | Saitual | Serchhip | Siaha | Mizoram |
| --- | --- | --- | --- | --- | --- | --- | --- | --- | --- | --- | --- | --- |
| OXK* |  |  |  |  |  |  |  |  |  |  |  |  |
| 2019 | 0.33 | 0.16 | 0.08 | 0.30 | 0.51 | 0.00 | 0.00 | 0.00 | 0.00 | 0.96 | 0.00 | 0.23 |
| 2020 | 0.89 | 0.29 | 0.11 | 1.26 | 0.11 | 0.00 | 0.00 | 1.90 | 0.66 | 0.16 | 0.11 | 0.57 |
| 2021 | 1.03 | 0.36 | 0.48 | 0.30 | 0.31 | 0.01 | 0.06 | 0.67 | 0.35 | 3.17 | 0.02 | 0.67 |
| 2022 | 2.00 | 1.04 | 12.36 | 9.66 | 2.49 | 0.95 | 0.59 | 5.77 | 7.78 | 9.39 | 0.17 | 3.04 |
| Avg. incidence rate | 1.06 | 0.46 | 3.26 | 2.88 | 0.86 | 0.24 | 0.16 | 2.08 | 2.20 | 3.42 | 0.07 | 1.13 |
| OX2 |  |  |  |  |  |  |  |  |  |  |  |  |
| 2019 | 0.01 | 0.42 | 2.91 | 2.60 | 0.09 | 0.00 | 0.00 | 0.00 | 0.00 | 0.00 | 0.04 | 0.20 |
| 2020 | 0 | 0.34 | 3.97 | 0.43 | 0.35 | 0 | 0.49 | 0.88 | 0 | 0 | 0.08 | 0.30 |
| 2021 | 0.17 | 0.29 | 1.12 | 0.16 | 0.33 | 0.00 | 0.41 | 0.11 | 0.68 | 0.17 | 0.00 | 0.24 |
| 2022 | 0.02 | 0.06 | 0.70 | 0.51 | 0.06 | 0.01 | 0.04 | 0.12 | 0.12 | 0.24 | 0.00 | 0.08 |
| Avg. incidence rate | 0.05 | 0.28 | 2.17 | 0.93 | 0.21 | 0.00 | 0.24 | 0.28 | 0.20 | 0.10 | 0.03 | 0.21 |
| OX19 |  |  |  |  |  |  |  |  |  |  |  |  |
| 2019 | 0.01 | 0.18 | 0.11 | 3.62 | 0.05 | 0.00 | 0.00 | 0.00 | 0.00 | 0.02 | 0.02 | 0.13 |
| 2020 | 0.00 | 0.08 | 0.08 | 2.95 | 0.06 | 0.00 | 0.01 | 0.54 | 0.00 | 0.00 | 0.08 | 0.15 |
| 2021 | 0.04 | 0.06 | 0.11 | 0.19 | 0.17 | 0.00 | 0.01 | 0.19 | 0.19 | 0.11 | 0.00 | 0.07 |
| 2022 | 0.02 | 0.06 | 0.39 | 0.56 | 0.11 | 0.03 | 0.02 | 0.31 | 0.17 | 0.17 | 0.00 | 0.10 |
| Avg. incidence rate | 0.02 | 0.09 | 0.17 | 1.83 | 0.10 | 0.01 | 0.01 | 0.26 | 0.09 | 0.07 | 0.03 | 0.11 |
| OXK and OX2 |  |  |  |  |  |  |  |  |  |  |  |  |
| 2019 | 0 | 0.03 | 0.08 | 0.56 | 0.11 | 0.00 | 0.00 | 0.00 | 0.00 | 0.00 | 0.83 | 0 |
| 2020 | 0.00 | 0.00 | 0.03 | 0.30 | 0.24 | 0.00 | 0.01 | 0.08 | 0.00 | 0.00 | 1.04 | 0.00 |
| 2021 | 0.01 | 0.05 | 0.06 | 0.00 | 0.12 | 0.00 | 0.00 | 0.01 | 0.03 | 0.02 | 0.64 | 0.01 |
| 2022 | 0.02 | 0.02 | 0.22 | 0.24 | 0.07 | 0.03 | 0.04 | 0.16 | 0.24 | 0.17 | 1.86 | 0.02 |
| Avg. incidence rate | 0.01 | 0.02 | 0.10 | 0.28 | 0.14 | 0.01 | 0.01 | 0.06 | 0.07 | 0.05 | 1.09 | 0.01 |
| OXK and OX19 |  |  |  |  |  |  |  |  |  |  |  |  |
| 2019 | 0.00 | 0.00 | 0.31 | 0.00 | 0.06 | 0.10 | 0.00 | 0.00 | 0.00 | 0.00 | 0.72 | 0.00 |
| 2020 | 0.04 | 0.00 | 0.14 | 0.27 | 0.18 | 0.32 | 0.00 | 0.02 | 0.00 | 0.00 | 2.16 | 0.04 |
| 2021 | 0.11 | 0.02 | 0.53 | 0.05 | 0.22 | 0.11 | 0.00 | 0.08 | 0.28 | 0.03 | 2.97 | 0.11 |
| 2022 | 0.03 | 0.04 | 1.29 | 0.59 | 0.11 | 0.04 | 0.06 | 0.31 | 0.35 | 0.44 | 4.03 | 0.03 |
| Avg. incidence rate | 0.05 | 0.01 | 0.57 | 0.23 | 0.14 | 0.15 | 0.01 | 0.10 | 0.16 | 0.12 | 2.47 | 0.05 |
| OX2 and OX19 |  |  |  |  |  |  |  |  |  |  |  |  |
| 2019 | 0 | 0.01 | 0.00 | 0.03 | 0.05 | 0.00 | 0.00 | 0.00 | 0.00 | 0.00 | 0.15 | 0 |
| 2020 | 0.00 | 0.01 | 0.03 | 0.21 | 0.04 | 0.00 | 0.00 | 0.14 | 0.00 | 0.00 | 0.61 | 0.00 |
| 2021 | 0.01 | 0.07 | 0.03 | 0.00 | 0.02 | 0.55 | 0.00 | 0.22 | 0.03 | 0.02 | 2.56 | 0.01 |
| 2022 | 0.06 | 0.07 | 0.39 | 0.32 | 0.11 | 0.14 | 0.04 | 0.36 | 0.23 | 0.41 | 3.69 | 0.06 |
| Avg. incidence rate | 0.02 | 0.04 | 0.11 | 0.14 | 0.05 | 0.17 | 0.01 | 0.18 | 0.07 | 0.11 | 1.75 | 0.02 |
| OXK, OX2 and OX19 |  |  |  |  |  |  |  |  |  |  |  |  |
| 2019 | 0.18 | 0.00 | 0.06 | 0.00 | 0.04 | 0.00 | 0.00 | 0.12 | 0.00 | 0.00 | 0.00 | 0.08 |
| 2020 | 0.43 | 0.02 | 0.00 | 0.03 | 0.05 | 0.00 | 0.00 | 0.12 | 0.00 | 0.00 | 0.00 | 0.17 |
| 2021 | 0.22 | 0.02 | 0.00 | 0.00 | 0.02 | 0.00 | 0.00 | 0.02 | 0.00 | 0.02 | 0.00 | 0.08 |
| 2022 | 0.16 | 0.04 | 0.25 | 0.54 | 0.01 | 0.05 | 0.04 | 0.40 | 0.26 | 0.35 | 0.02 | 0.15 |
| Avg. incidence rate | 0.25 | 0.02 | 0.08 | 0.14 | 0.03 | 0.01 | 0.01 | 0.17 | 0.07 | 0.09 | 0.01 | 0.12 |

*In 2018, only two OXK positive cases were reported and hence not included.
