## Supplementary Table 5 for "Epidemiology of scrub typhus and other rickettsial infections (2018-22) in the hyper-endemic setting of Mizoram, North-East India"

**Supplementary Table 5 Age and gender based distribution of rickettsial infections in Mizoram (2018-2022)**

| Age groups (in years) | | Scrub typhus | Other rickettsial infections | | | Mixed infections | | | Total |
| --- | --- | --- | --- | --- | --- | --- | --- | --- | --- |
|  |  |  | **OX2** | **OX19** | **OX2 and OX19** | **OXK and OX2** | **OXK and OX19** | **OXK, OX2 and OX 19** |  |
| <1 year |  |  |  |  |  |  |  |  |  |
|  | Male | 45 | 5 | 1 | 2 | 1 | 2 | 0 | 56 |
|  | Female | 33 | 1 | 0 | 0 | 1 | 2 | 0 | 37 |
|  | Total | 78 (0.40%) | 6 (0.56%) | 1 (0.17%) | 2 (0.97%) | 2 (0.43%) | 4 (1.21%) | 0 | 93 (0.40%) |
| 1-5 |  |  |  |  |  |  |  |  |  |
|  | Male | 576 | 10 | 13 | 5 | 11 | 10 | 19 | 644 |
|  | Female | 520 | 6 | 13 | 9 | 2 | 13 | 9 | 572 |
|  | Total | 1096 (5.58%) | 16 (1.50%) | 26 (4.55%) | 14 (6.8%) | 13 (2.79%) | 23 (6.95%) | 28 (4.52%) | 1216 (5.30%) |
| 6-10 |  |  |  |  |  |  |  |  |  |
|  | Male | 688 | 18 | 27 | 8 | 7 | 14 | 28 | 790 |
|  | Female | 512 | 15 | 29 | 6 | 9 | 6 | 6 | 583 |
|  | Total | 1200 (6.11%) | 33 (3.09%) | 56 (9.79%) | 14 (6.8%) | 16 (3.43%) | 20 (6.04%) | 34 (5.48%) | 1373 (5.99%) |
| 11-20 |  |  |  |  |  |  |  |  |  |
|  | Male | 1106 | 29 | 36 | 12 | 21 | 16 | 36 | 1256 |
|  | Female | 844 | 32 | 41 | 14 | 15 | 20 | 27 | 993 |
|  | Total | 1950 (9.92%) | 61 (5.71%) | 77 (13.46%) | 26 (12.62%) | 36 (7.73%) | 36 (10.88%) | 63 (10.16%) | 2249 (9.81%) |
| 21-30 |  |  |  |  |  |  |  |  |  |
|  | Male | 1580 | 66 | 44 | 16 | 29 | 18 | 43 | 1796 |
|  | Female | 1300 | 57 | 65 | 10 | 34 | 31 | 34 | 1531 |
|  | Total | 2880 (14.66%) | 123 (11.52%) | 109 (19.06%) | 26 (12.62%) | 63 (13.52%) | 49 (14.80%) | 77 (12.42%) | 3327 (14.51%) |
| 31-40 |  |  |  |  |  |  |  |  |  |
|  | Male | 1803 | 94 | 38 | 19 | 39 | 28 | 66 | 2087 |
|  | Female | 1676 | 98 | 61 | 21 | 29 | 35 | 57 | 1977 |
|  | Total | 3479 (17.70%) | 192 (17.98%) | 99 (17.31%) | 40 (19.42%) | 68 (14.59%) | 63 (19.03%) | 123 (19.84%) | 4064 (17.73%) |
| 41-50 |  |  |  |  |  |  |  |  |  |
|  | Male | 1671 | 95 | 36 | 10 | 33 | 23 | 59 | 1927 |
|  | Female | 1581 | 127 | 67 | 7 | 30 | 24 | 61 | 1897 |
|  | Total | 3252 (16.55%) | 222 (20.79%) | 103 (18.01%) | 17 (8.25%) | 63 (13.52%) | 47 (14.2%) | 120 (19.35%) | 3824 (16.68%) |
| 51-60 |  |  |  |  |  |  |  |  |  |
|  | Male | 1228 | 94 | 23 | 17 | 32 | 11 | 31 | 1436 |
|  | Female | 1357 | 86 | 23 | 17 | 44 | 23 | 44 | 1594 |
|  | Total | 2585 (13.15%) | 180 (16.85%) | 46 (8.04%) | 34 (16.5%) | 76 (16.31%) | 34 (10.27%) | 75 (12.10%) | 3030 (13.22%) |
| 61-70 |  |  |  |  |  |  |  |  |  |
|  | Male | 935 | 57 | 18 | 10 | 34 | 17 | 37 | 1109 |
|  | Female | 901 | 55 | 11 | 7 | 50 | 16 | 21 | 1061 |
|  | Total | 1836 (9.34%) | 112 (10.49%) | 29 (5.07%) | 17 (8.25%) | 84 (18.03%) | 33 (9.97%) | 59 (9.52%) | 2170 (9.47%) |
| 71-80 |  |  |  |  |  |  |  |  |  |
|  | Male | 493 | 47 | 10 | 6 | 20 | 4 | 21 | 600 |
|  | Female | 485 | 47 | 7 | 8 | 18 | 14 | 12 | 591 |
|  | Total | 978 (4.98%) | 94 (8.80%) | 17 (2.97%) | 14 (6.8%) | 38 (8.15%) | 18 (5.44%) | 32 (5.16%) | 1191 (5.19%) |
| 81-90 |  |  |  |  |  |  |  |  |  |
|  | Male | 140 | 15 | 4 | 1 | 1 | 4 | 4 | 169 |
|  | Female | 136 | 10 | 5 | 1 | 4 | 0 | 3 | 159 |
|  | Total | 276 (1.40%) | 25 (2.34%) | 9 (1.57%) | 2 (0.97%) | 5 (1.07%) | 4 (1.21%) | 7 (1.13%) | 328 (1.43%) |
| 91 and above |  |  |  |  |  |  |  |  |  |
|  | Male | 13 | 4 | 0 | 0 | 0 | 0 | 2 | 19 |
|  | Female | 28 | 0 | 0 | 0 | 2 | 0 | 0 | 30 |
|  | Total | 41 (0.21%) | 4 (0.37%) | 0 (0%) | 0 (0%) | 2 (0.43%) | 0 (0%) | 2 (0.32%) | 49 (0.18%) |
| Cumulative case count | Male  Female  Total | 10278 (52.3%)  9373 (47.7%)  19651 | 534 (50%)  534 (50%)  1068 | 250 (43.7%)  322 (56.3%)  572 | 106 (51.5%)  100 (48.5%)  206 | 228 (48.9%)  238 (51.1%)  466 | 147 (44.4%)  184 (55.6%)  331 | 346 (55.8%)  274 (44.2%)  620 | 11889 (51.89%)  11025 (48.11%)  22914 |
