## Supplementary Table 6 for "Epidemiology of scrub typhus and other rickettsial infections (2018-22) in the hyper-endemic setting of Mizoram, North-East India"

**Supplementary Table 6.** **Distribution of rickettsial cases across different occupational groups in Mizoram (2018-2022)**

| Occupation | Scrub typhus | | Other infections | | | | | | Mixed infections | | | | | | Total | (%) |
| --- | --- | --- | --- | --- | --- | --- | --- | --- | --- | --- | --- | --- | --- | --- | --- | --- |
|  |  |  | **OX2** | | **OX19** | | **OX2 and OX19** | | **OXK and OX2** | | **OXK and OX19** | | **OXK, OX2 and OX 19** | |  |  |
|  | Cases | (%) | Cases | (%) | Cases | (%) | Cases | (%) | Cases | (%) | Cases | (%) | Cases | (%) |  |  |
| Business | 1955 | 9.9 | 64 | 6.0 | 23 | 4.0 | 22 | 10.7 | 38 | 8.2 | 23 | 6.9 | 63 | 10.2 | 2188 | 9.5 |
| Construction workers | 1104 | 5.6 | 94 | 8.8 | 9 | 1.6 | 8 | 3.9 | 28 | 6.0 | 13 | 3.9 | 29 | 4.7 | 1285 | 5.6 |
| Farmer | 9805 | 49.9 | 672 | 62.9 | 283 | 49.5 | 93 | 45.1 | 282 | 60.5 | 171 | 51.7 | 318 | 51.3 | 11624 | 50.7 |
| Government services | 1344 | 6.8 | 56 | 5.2 | 44 | 7.7 | 15 | 7.3 | 22 | 4.7 | 15 | 4.5 | 54 | 8.7 | 1550 | 6.8 |
| Pre school | 968 | 4.9 | 16 | 1.5 | 18 | 3.1 | 13 | 6.3 | 13 | 2.8 | 25 | 7.6 | 26 | 4.2 | 1079 | 4.7 |
| Students | 4301 | 21.9 | 155 | 14.5 | 193 | 33.7 | 54 | 26.2 | 76 | 16.3 | 78 | 23.6 | 125 | 20.2 | 4982 | 21.7 |
| Others (aged 71 and above) | 174 | 0.9 | 11 | 1.0 | 2 | 0.3 | 1 | 0.5 | 7 | 1.5 | 6 | 1.8 | 5 | 0.8 | 206 | 0.9 |
|  | **19651** |  | **1068** |  | **572** |  | **206** |  | **466** |  | **331** |  | **620** |  | **22914** |  |
