## Supplementary Table 7 for "Epidemiology of scrub typhus and other rickettsial infections (2018-22) in the hyper-endemic setting of Mizoram, North-East India"

**Supplementary Table 7 Summarized table of clinical presentations of the reported rickettsial infections in Mizoram**

| Clinical presentations | Scrub typhus | Other Rickettsial infections | | | | | Mixed infections | | | | | Total (%) |
| --- | --- | --- | --- | --- | --- | --- | --- | --- | --- | --- | --- | --- |
|  |  | **OX2 Reactive** | **OX19 Reactive** | | **OX19 &OX2 Reactive** | | **Scrub typhus (OXK) & OX19 Reactive** | **Scrub typhus (OXK) &OX2 Reactive** | | **Scrub typhus (OXK), OX2 &OX19 Reactive** | |  |
| Fever/  Persistent fever | 16832 | 821 | | 469 | | 156 | 250 | | 408 | | 547 | 19483 (85.03) |
| Bodyache | 1703 | 58 | | 32 | | 16 | 43 | | 56 | | 57 | 1965 (8.58) |
| Headache | 5719 | 331 | | 175 | | 70 | 128 | | 157 | | 146 | 6726 (29.35) |
| Chills | 4237 | 197 | | 139 | | 49 | 55 | | 68 | | 150 | 4895 (21.36) |
| Cough | 1851 | 69 | | 48 | | 19 | 27 | | 33 | | 66 | 2113 (9.22) |
| Rash | 5315 | 274 | | 197 | | **56** | 68 | | 89 | | 141 | 6140 (26.80) |
| Eschar | 443 | **76** | | 9 | | 7 | 7 | | 29 | | 10 | 581 (2.54) |
| Vomiting | 411 | 35 | | 17 | | 7 | 8 | | 7 | | 5 | 490 (2.14) |
| CNS involvement | 377 | 20 | | 16 | | 4 | 5 | | 5 | | 14 | 441 (1.92) |
| Nausea | 634 | 23 | | 21 | | 12 | 14 | | 10 | | 26 | 740 (3.23) |
| Loose stool/  loose motion | 29 | 0 | | 0 | | 0 | 0 | | 0 | | 1 | 30 (0.13) |
| Others* | 24 | 1 | | 0 | | 0 | 0 | | 1 | | 0 | 26 (0.11) |

*Others: Abdominal pain, enlarged lymph nodes, ascites, weakness, breathing difficulty/shortness of breath, lethargy
