## Supplementary Table 8 for "Epidemiology of scrub typhus and other rickettsial infections (2018-22) in the hyper-endemic setting of Mizoram, North-East India"

**Supplementary Table 8 Distribution of clinical presentations among different occupational groups in Mizoram**

|  | Farmer | Business | Construction worker | Govt. services | Others  (aged 71 and above) | Pre school | Student | Total | (%) |
| --- | --- | --- | --- | --- | --- | --- | --- | --- | --- |
| Fever/  Persistent fever | 9924 | 1839 | 1108 | 1328 | 163 | 921 | 4200 | 19483 | 85.03 |
| Bodyache | 978 | 190 | 127 | 135 | 21 | 89 | 425 | 1965 | 8.58 |
| Headache | 3429 | 602 | 355 | 454 | 76 | 306 | 1504 | 6726 | 29.35 |
| Chills | 2425 | 505 | 228 | 332 | 49 | 270 | 1086 | 4895 | 21.36 |
| Cough | 968 | 240 | 113 | 162 | 16 | 127 | 487 | 2113 | 9.22 |
| Rash | 3028 | 631 | 290 | 416 | 60 | 304 | 1411 | 6140 | 26.80 |
| Vomiting | 245 | 43 | 21 | 29 | 6 | 26 | 120 | 490 | 2.14 |
| CNS involvement | 235 | 43 | 14 | 27 | 4 | 19 | 99 | 441 | 1.92 |
| Loose stool/  loose motion | 8 | 8 | 6 | 0 | 0 | 1 | 7 | 30 | 0.13 |
| Nausea | 359 | 82 | 32 | 56 | 8 | 42 | 161 | 740 | 3.23 |
| Eschar | 306 | 53 | 41 | 40 | 12 | 16 | 113 | 581 | 2.54 |
| Others* | 14 | 4 | 2 | 4 | 0 | 1 | 1 | 26 | 0.11 |

*Others: Abdominal pain, enlarged lymph nodes, ascites, weakness, breathing difficulty/shortness of breath, lethargy
