## Supplementary Table 9 for "Epidemiology of scrub typhus and other rickettsial infections (2018-22) in the hyper-endemic setting of Mizoram, North-East India"

**Supplementary Table 9 Distribution of hospitalized rickettsial and scrub typhus cases in Mizoram (2018-2022)**

|  | Rickettsial infections | | Scrub typhus cases | |
| --- | --- | --- | --- | --- |
|  | **Hospitalized** | **Not hospitalized** | **Hospitalized** | **Not hospitalized** |
|  | **4182 (18.25 %)** | **18732 (81.75 %)** | **3832 (19.50 %)** | **15819 (80.45 %)** |
| Age (in years) |  |  |  |  |
| <1 year | 15 (16.13) | 78 (83.87) | 13 (16.67) | 65 (83.33) |
| 1 to 5 | 251 (20.64) | 965 (79.36) | 234 (21.35) | 862 (78.65) |
| 6 to 10 | 292 (21.27) | 1081 (78.73) | 270 (22.50) | 930 (77.50) |
| 11 to 20 | 403 (17.92) | 1846 (82.08) | 375 (19.23) | 1575 (80.77) |
| 21 to 30 | 639 (19.21) | 2688 (80.79) | 591 (20.52) | 2289 (79.48) |
| 31 to 40 | 682 (16.78) | 3382 (83.22) | 635 (18.25) | 2844 (81.75) |
| 41 to 50 | 607 (15.87) | 3217 (84.13) | 564 (17.34) | 2688 (82.66) |
| 51 to 60 | 510 (16.83) | 2520 (83.17) | 474 (18.34) | 2111 (81.66) |
| 61 to 70 | 388 (17.88) | 1782 (82.12) | 348 (18.95) | 1488 (81.05) |
| 71 to 80 | 287 24.10) | 904 (75.90) | 237 (24.23) | 741 (75.77) |
| 81 to 90 | 88( (26.83) | 240 (73.17) | 75 (27.17) | 201 (72.83) |
| 91 and above | 20 (40.82) | 29 (59.18) | 16 (39.02) | 25 (60.98) |
| Sex |  |  |  |  |
| Female | 2055 (18.64) | 8970 (81.36) | 1866 (19.91) | 7507 (80.09) |
| Male | 2127 (17.89) | 9762 (82.11) | 1966 (19.13) | 8312 (80.87) |
| Occupation |  |  |  |  |
| Business | 440 (20.11) | 1748 (79.89) | 405 (20.72) | 1550 (79.28) |
| Construction workers | 132 (10.27) | 1153 (89.73) | 114 (10.33) | 990 (89.67) |
| Farmer | 1898 (16.33) | 9726 (83.67) | 1725 (17.59) | 8080 (82.41) |
| Government services | 298 (19.23) | 1252 (80.77) | 278 (20.68) | 1066 (79.32) |
| Pre school | 217 (20.11) | 862 (79.89) | 200 (20.66) | 768 (79.34) |
| Students | 1068 (21.44) | 3914 (78.56) | 999 (23.23) | 3302 (76.77) |
| Others (aged 71 and above) | 129 (62.62) | 77 (37.38) | 111 (63.79) | 63 (36.21) |
| Deaths | 62 (76.54) | 19 (23.46) | 23(30.67) | 52 (69.33) |
