## Supplementary Table 10 for "Epidemiology of scrub typhus and other rickettsial infections (2018-22) in the hyper-endemic setting of Mizoram, North-East India"

**Supplementary Table 10 Case fatality rates (CFRs) of different rickettsial infections in the state of Mizoram, 2018-2022**

| Variables | Scrub typhus | | | Other rickettsial infections | | | Mixed infections | | | Total rickettsial deaths |
| --- | --- | --- | --- | --- | --- | --- | --- | --- | --- | --- |
|  | **Cases** | **Deaths** | **CFR** | **Cases** | **Deaths** | **CFR** | **Cases** | **Deaths** | **CFR** | **n (%)** |
| Age |  |  |  |  |  |  |  |  |  |  |
| Less than 11 years | 2374 | 12 | 0.51 | 168 | 1 | 0.60 | 140 | 1 | 0.71 | 14 (17.28) |
| 11-30 years | 4830 | 14 | 0.29 | 422 | 0 | 0.00 | 324 | 0 | 0.00 | 14 (17.28) |
| 31-50 years | 6731 | 16 | 0.24 | 673 | 1 | 0.15 | 484 | 1 | 0.21 | 18 (22.22) |
| 51-70 years | 4421 | 29 | 0.66 | 418 | 0 | 0.00 | 361 | 1 | 0.28 | 30 (37.04) |
| 71 and above | 1295 | 4 | 0.31 | 165 | 0 | 0.00 | 108 | 1 | 0.93 | 5 (6.17) |
| Sex |  |  |  |  |  |  |  |  |  |  |
| Male | 10278 | 48 | 0.47 | 890 | 1 | 0.11 | 721 | 2 | 0.28 | 51 (62.96) |
| Female | 9373 | 27 | 0.29 | 956 | 1 | 0.10 | 696 | 2 | 0.29 | 30 (37.04) |
| Districts |  |  |  |  |  |  |  |  |  |  |
| Aizawl | 9863 | 50 | 0.51 | 149 | 0 | 0.00 | 568 | 2 | 0.35 | 52 964.20) |
| Champhai | 407 | 2 | 0.49 | 164 | 2 | 1.22 | 30 | 0 | 0.00 | 4 (4.94) |
| Hnahthial | 531 | 0 | 0.00 | 350 | 0 | 0.00 | 108 | 1 | 0.93 | 1 (1.23) |
| Khawzawl | 453 | 0 | 0.00 | 452 | 0 | 0.00 | 76 | 0 | 0.00 | 0 (0.00) |
| Kolasib | 459 | 2 | 0.44 | 191 | 0 | 0.00 | 97 | 1 | 1.03 | 3 (3.70) |
| Lawngtlai | 1357 | 9 | 0.66 | 10 | 0 | 0.00 | 196 | 0 | 0.00 | 9 (11.11) |
| Lunglei | 1125 | 2 | 0.18 | 143 | 0 | 0.00 | 19 | 0 | 0.00 | 2 (2.47) |
| Mamit | 1082 | 5 | 0.46 | 236 | 0 | 0.00 | 176 | 0 | 0.00 | 5 (6.17) |
| Saitual | 894 | 1 | 0.11 | 83 | 0 | 0.00 | 66 | 0 | 0.00 | 1 (1.23) |
| Serchhip | 2975 | 4 | 0.13 | 57 | 0 | 0.00 | 80 | 0 | 0.00 | 4 (4.94) |
| Siaha | 505 | 0 | 0.00 | 11 | 0 | 0.00 | 1 | 0 | 0.00 | 0 (0.00) |
| Occupation |  |  |  |  |  |  |  |  |  |  |
| Farmer | 9805 | 31 | 0.32 | 1047 | 1 | 0.10 | 771 | 3 | 0.39 | 35 (43.21) |
| Business | 1955 | 12 | 0.61 | 109 | 0 | 0.00 | 124 | 0 | 0.00 | 12 (14.81) |
| Construction worker | 1104 | 13 | 1.18 | 111 | 0 | 0.00 | 70 | 0 | 0.00 | 13 (16.05) |
| Government Services | 1344 | 2 | 0.15 | 115 | 0 | 0.00 | 91 | 0 | 0.00 | 2 (2.47) |
| Preschool | 968 | 4 | 0.41 | 47 | 0 | 0.00 | 64 | 0 | 0.00 | 4 (4.94) |
| Student | 4301 | 12 | 0.28 | 401 | 1 | 0.25 | 401 | 1 | 0.36 | 14 (17.28) |
| Others (aged 71 and above) | 174 | 1 | 0.57 | 14 | 0 | 0.00 | 14 | 0 | 0.00 | 1 (1.23) |
| Eschar |  |  |  |  |  |  |  |  |  |  |
| Yes | 443 | 5 | 1.13 | 92 | 0 | 0.00 | 46 | 0 | 0.00 | 5 (6.17) |
| No | 19208 | 70 | 0.36 | 1752 | 2 | 0.11 | 1371 | 4 | 0.29 | 76 (93.83) |
| Hospitalization |  |  |  |  |  |  |  |  |  |  |
| Yes | 3832 | 56 | 1.16 | 146 | 2 | 1.37 | 202 | 4 | 1.98 | 62 (76.54) |
| No | 15819 | 19 | 0.12 | 1698 | 2 | 0.12 | 1215 | 0 | 0.00 | 19 (23.46) |
| Year |  |  |  |  |  |  |  |  |  |  |
| 2018 | 2183 | 6 | 0.27 |  |  |  |  |  |  | 6 (7.41) |
| 2019 | 5859 | 8 | 0.14 | 472 | 1 | 0.21 | 141 | 0 | 0.00 | 9 (11.11) |
| 2020 | 2744 | 14 | 0.51 | 626 | 1 | 0.16 | 347 | 3 | 0.86 | 18 (22.22) |
| 2021 | 2323 | 21 | 0.90 | 430 | 0 | 0.00 | 367 | 0 | 0.00 | 21 (25.93) |
| 2022 | 6542 | 26 | 0.40 | 318 | 0 | 0.00 | 562 | 1 | 0.18 | 27 (33.33) |
| 2018-2022 | **19651** | **75** | **0.38** | **1846** | **2** | **0.11** | **1417** | **4** | **0.28** | **81 (100)** |
