## Supplementary Table 11 for "Epidemiology of scrub typhus and other rickettsial infections (2018-22) in the hyper-endemic setting of Mizoram, North-East India"

**Supplementary Table 11. Association between eschar and scrub typhus disease outcome**

|  |  |  | **Scrub typhus outcome** | | **p-value** |
| --- | --- | --- | --- | --- | --- |
|  |  |  | **Recovered** | **Deaths** |  |
| **Eschar** | **Yes** | Cases | 438 | 5 | 0.010 |
|  |  | Expected | 441.3 | 1.7 |  |
|  | **No** | Cases | 19138 | 70 |  |
|  |  | Expected | 19134.7 | 73.3 |  |

p<0.05 is significant
