## Supplementary figures and images for "Epidemiology of scrub typhus and other rickettsial infections (2018-22) in the hyper-endemic setting of Mizoram, North-East India"

### Supplementary Figure 1

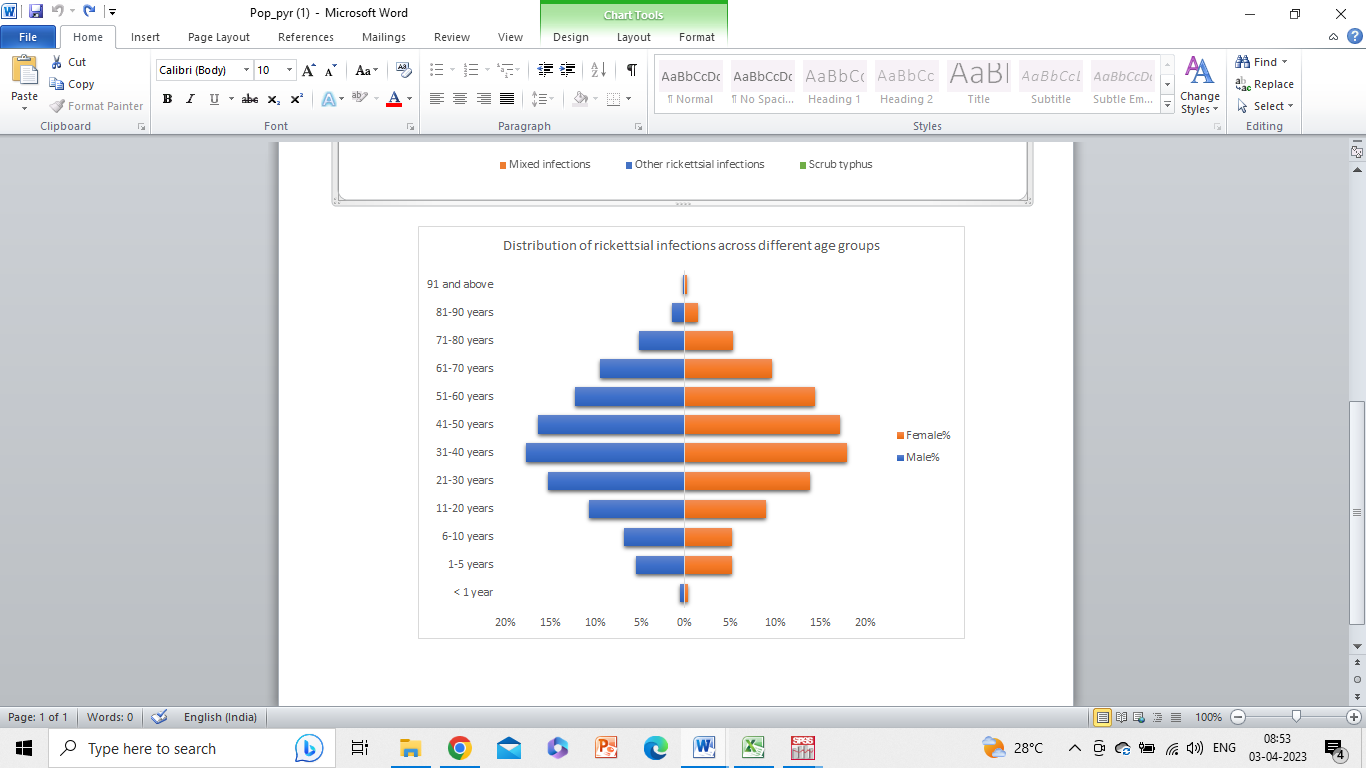
**Supplementary Figure 1.** **Distribution of rickettsial infections across different age groups**
